# Genetic effects on serum and urine metabolites across ancestries and their links to disease

**DOI:** 10.64898/2026.11.09.26367601

**Authors:** Inga Steinbrenner, Ruiyuan Zhang, Nora Scherer, Yura Lee, Ngoc Quynh H Nguyen, Alex Waterhölter, Zulema Rodriguez-Hernandez, Oleg Borisov, Dmitrii Zakharenko, Aditya Surapaneni, Sara Monteiro-Martins, Yong Li, Tanika N. Kelly, Jing Chen, Jiang He, Anna Köttgen, Eric Boerwinkle, Josef Coresh, Morgan E. Grams, Changwei Li, Bing Yu, Pascal Schlosser

**Author notes:** Corresponding authors: Bing Yu, PhD, Department of Epidemiology School of Public Health, The University of Texas Health Science Center at Houston Houston, Texas, USA, Pascal Schlosser, PhD, Institute of Epidemiology and Prevention Medical Center – University of Freiburg Hugstetter Str. 49, 79106 Freiburg, Germany. Equal contribution (Inga Steinbrenner, Ruiyuan Zhang, Bing Yu, Pascal Schlosser).

## Abstract

Genome-wide association studies of the metabolome have predominantly examined European-ancestry populations using a single biofluid, constraining both the discovery of metabolite quantitative trait loci (mQTLs) and the resolution of causal variant finemapping. Here, we report a multi-ancestry genome-wide association study of 1,955 serum and urine metabolites in 4,431 European American and African American participants from the Atherosclerosis Risk in Communities study. We identify 969 mQTLs for 553 metabolites, 30% unreported in studies of matched biofluid and ancestry, and replicate 91-99% of testable mQTLs in two independent cohorts. In the African American sample with balanced sample sizes, 36% of metabolites with mQTLs are serum-specific, 39% urine-specific, and 25% shared, demonstrating complementary genetic architecture. Only 23 of 632 candidate ancestry-specific mQTLs show heterogeneous effects beyond variant availability and power, yet allele frequency differences expose functional variants such as an *SLC28A1* splice variant, virtually absent in Europeans, that increases urine uridine excretion, implicating impaired CNT1-mediated renal nucleoside reabsorption. Multi-ancestry finemapping improves credible set resolution, and colocalization with the Million Veteran Program phenome links the catalog to cardiometabolic, kidney and liver traits consistent with prioritized gene function. Integrating biofluids and ancestries enhances genetic discovery and mechanistic characterization of the human metabolome.

## Introduction

Genome-wide association studies of the metabolome (mGWAS) connect genetic variation to small-molecule levels, and thereby to the enzymes and transporters that produce, convert and clear them. Targeted nuclear magnetic resonance (NMR) panels such as Nightingale quantify about 250 predominantly lipoprotein, lipid, fatty-acid, amino-acid and glycolysis-related traits in up to 619,372 individuals.^1,2^ Untargeted mass-spectrometry panels such as Metabolon resolve more than a thousand molecules, including amino acids, lipids, nucleotides, xenobiotics, and unnamed compounds.^3–7^

The strongest signals typically fall in genes whose products act directly on the associated metabolite,^8,9^ such as the desaturase *FADS1* for polyunsaturated phospholipids, and the transporter *SLC16A9* for carnitine and urate^10^. Because such loci often also associate with disease, metabolites can point to candidate causal genes at disease loci and pose a mechanistic hypothesis for experimental validation.

Joint profiling of cerebrospinal fluid (CSF) and plasma proteomes has led to major advances^11^ in neurological disease, and CSF metabolomics has proven similarly complementary to blood.^12^ For metabolomics, most efforts have focused on serum or plasma,^1,8,13^ leaving the urine metabolome, which reflects renal handling and xenobiotic clearance, under-investigated despite a growing number of efforts.^7,14^ Paired plasma and urine mGWAS have revealed enzymatic and transport processes at their interface, motivating joint modeling.^14^ Moreover, the metabolome has been mapped in only a few multi-ancestry mGWAS.^1,15,16^ Multi-ancestry designs exploit differences in allele frequency and linkage disequilibrium (LD) to increase power and finemapping resolution, as the Million Veteran Program (MVP) showed for both variant discovery and credible set resolution.^17^ Participants of African American as well as African ancestry remain severely underrepresented in GWAS and mGWAS efforts to date with few exceptions^18–20^, despite the greater genetic diversity and shorter LD blocks on average in African American ancestry populations, and no Metabolon-based urine mGWAS in African American participants has been reported. Ancestrally diverse reference panels such as TOPMed have improved imputation accuracy in these populations.^21,22^ They also recover small insertions and deletions (indels), a variant class that earlier panels captured poorly^23^ and that has therefore been largely absent from previous mGWAS. Here, we mapped the genetic regulation of 1,955 serum and urine metabolites in European American (EA) and African American (AA) participants of the Atherosclerosis Risk in Communities (ARIC) study^24,25^, using Metabolon profiling and TOPMed-based imputation. We asked what each biofluid contributes, whether ancestry-differentiated signals reflect biology or allele frequency, which enzymes and transporters multi-ancestry finemapping prioritizes, and how the catalog connects to disease.

## Results

### A catalog of 969 mQTLs across two biofluids and two ancestries

We conducted a multi-ancestry mGWAS of the serum and urine metabolome in participants of EA and AA ancestry with available genotype data and metabolomic profiling from visit 5 (2011–2013) from the ARIC study (serum: 3,607 EA / 818 AA; urine: 797 EA / 794 AA; both biofluids: 1,585; **Supplementary Table 1**; **Extended Data Fig. 1**). The participants were on average 76 years old, with a mean estimated glomerular filtration rate (eGFR) of 72 mL/min/1.73m² (serum) and 66 mL/min/1.73m² (urine), and clinical characteristics were comparable across ancestry-by-biofluid strata (**Supplementary Table 1**). After stringent quality control, 1,316 serum and 1,374 urine metabolites, harmonized to 1,955 unique analytes, were analyzed within each stratum (Methods; **Supplementary Table 2**). Genomic inflation factors were well-controlled across all GWAS (λ range: 0.98–1.02; median = 1.001; mean = 1.001) and within each ancestry-by-biofluid stratum (EA serum: mean λ = 1.005; EA urine: 1.002; AA serum: 1.000; AA urine: 0.999; **Supplementary Table 2**), arguing against residual population stratification.

A total of 969 unique metabolite quantitative trait loci (mQTLs) were identified across all analyses (*P* < 5 × 10⁻⁸ / 1,955; **Supplementary Tables 3-4, Extended Data Fig. 2**). Each mQTL is a metabolite-by-locus association, with up to four mQTLs per metabolite within an ancestry-biofluid stratum (**Fig. 1**; Methods); because the 1 Mb window merges rather than splits nearby signals, this definition is conservative with respect to locus counts. The 969 mQTLs involve 553 unique metabolites, fall into 287 non-overlapping genomic regions and were assigned to 247 most likely underlying genes (**Supplementary Table 5**). Among EA participants, we identified 620 serum mQTLs for 470 metabolites and 138 urine mQTLs for 130 metabolites, and among AA participants 102 serum mQTLs for 97 metabolites and 109 urine mQTLs for 101 metabolites. Effect directions were consistent in the other biofluid for 88% (EA) and 90% (AA) of mQTLs measured in both (**Supplementary Tables 3-4**). Notably, 58 (6%) of the index variants were small insertions and deletions, reflecting the enhanced variant resolution of TOPMed-based imputation. The index variant explained a median of 10.7% of metabolite variance (Q1-Q3: 6.6–15.7%). Higher values in urine than in serum (median 14.8% vs. 8.8%) and in AA than in EA (14.4% vs. 9.2%) reflected winner’s curse in the smaller strata rather than a difference in architecture: in AA, where serum and urine sample sizes are closely matched, the urine-versus-serum difference largely disappeared (14.5% vs. 14.2%; **Methods**).

**Figure 1:**
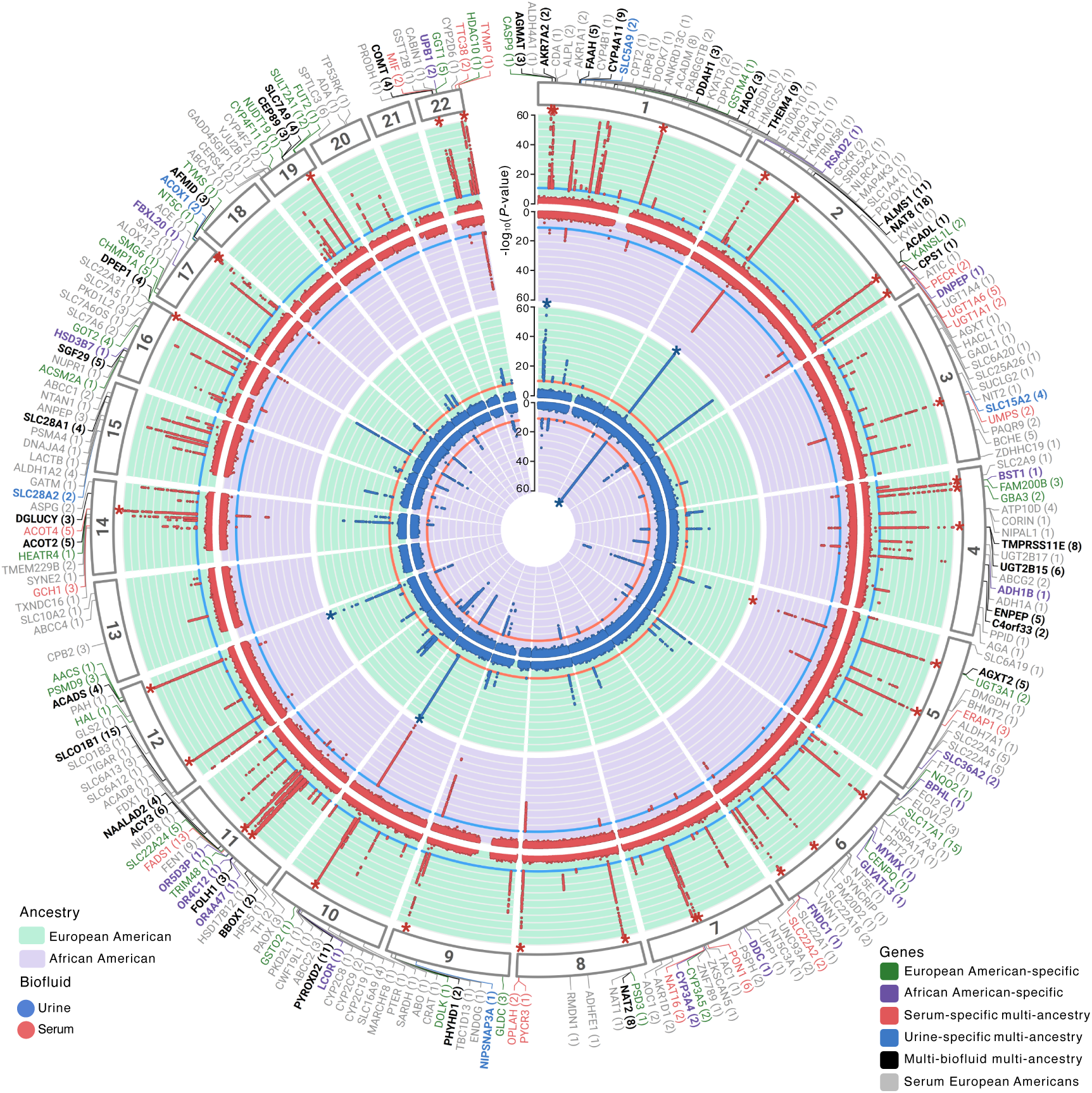
Genome-wide representation of metabolite quantitative trait loci (mQTLs). Outer ring: chromosomal position and prioritized gene (metabolite count in parentheses). Gene labels are colored by candidate specificity category: European American (EA)-specific (green), African American (AA)-specific (purple), serum-specific multi-ancestry (red), urine-specific multi-ancestry (blue), multi-biofluid multi-ancestry (black), and serum EA-only (gray). Inner Manhattan tracks show −log₁₀(*P*-value) values for EA (light green background) and AA (light purple background) associations, with serum (red dots) and urine (blue dots) mQTLs. Stars denote associations capped at *P* < 10^-60^. The inner red/blue lines indicate genome-wide significance threshold.

### Serum and urine capture complementary genetic regulation

Of the 1,955 unique metabolites 581 were only detected in serum, 639 only in urine, and 735 in both biofluids with overall aligned effect sizes across biofluids in both ancestries (**Fig. 2a,b**). In the AA cohort, where we have comparable serum and urine sample sizes (N_serum_ = 818, N_urine_ = 794), 211 mQTLs were detected for 159 unique metabolites of which 58 (36%) only showed associations with their levels in serum, 62 (39%) only in urine and 39 (25%) showed associations in both biofluids (**Fig. 3a, Supplementary Tables** 3-**4**; EA: 67 metabolites with mQTLs in both biofluids, **Extended Data Fig. 3**). Among the metabolites measured in both biofluids, 41 exhibited biofluid-specific mQTLs, which would have been missed if only one biofluid had been analyzed. This aligns with the proportions previously reported for an EA cohort,^7^ highlighting the specific value of urinary metabolite quantification and, more broadly, the importance of conducting multi-biofluid molecular GWAS.^14^ The increased statistical power from larger serum sample size in EA shifted proportions accordingly (76% serum only, 12% urine only, 13% both; **Extended Data Fig. 3**).

**Figure 2:**
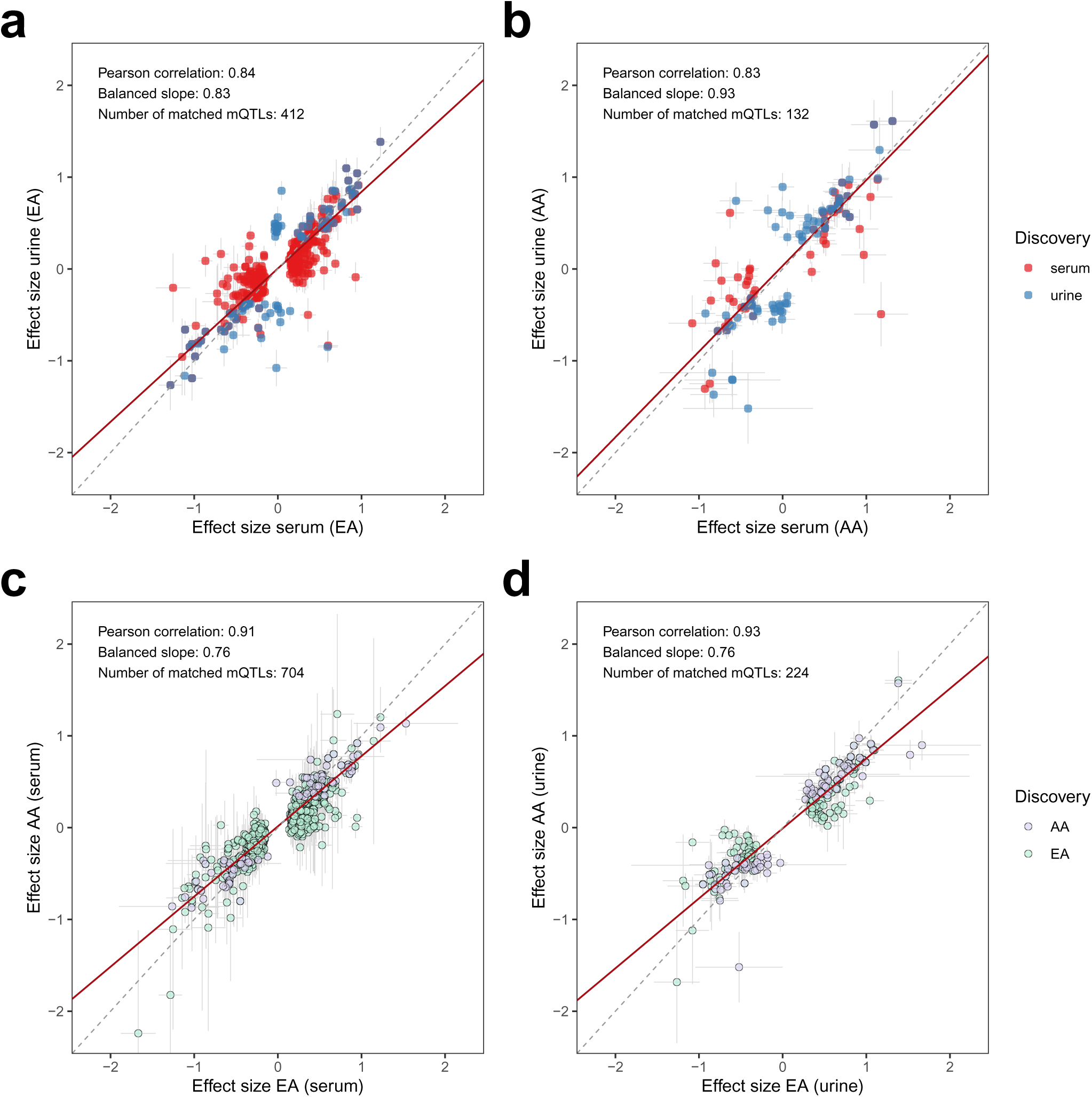
Combined comparison of index variant effect sizes across ancestries and biofluids in ARIC. Each point represents an mQTL, with error bars indicating ±1.96 × standard error. The dashed gray line represents the identity line (slope = 1). Pearson correlation coefficients, balanced slopes (red line, **Methods**), and the total number of matched mQTLs are annotated in each panel. Cross-biofluid comparison of EA (a; N=758) and AA (b, N=211) mQTLs, with serum effect sizes on the x-axis and urine effect sizes on the y-axis. Points are colored by discovery biofluid (red: serum, blue: urine). Cross-ancestry comparison of serum (c, N=722) and urine (d, N=247) mQTLs, with EA effect sizes on the x-axis and AA effect sizes on the y-axis. Points are colored by discovery ancestry (green: EA, purple: AA).

**Figure 3:**
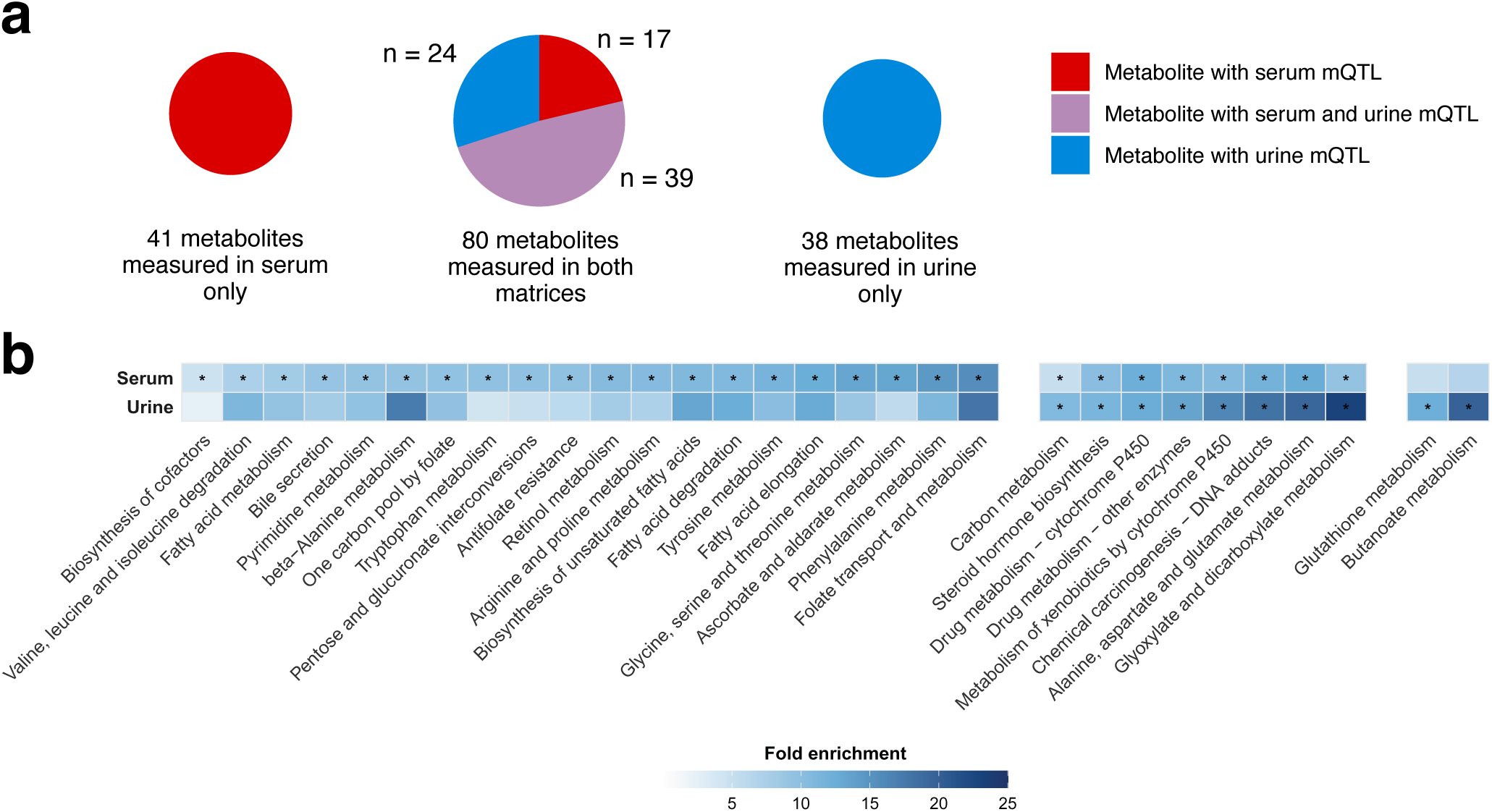
Characterization of metabolite quantitative trait loci (mQTL) in serum and urine. **(a)** Overview of African American (AA) mQTLs across biofluids. Pie charts show AA metabolites with mQTLs measured only in serum (left), only in urine (right), or in both (center), subdivided into those with serum mQTLs only (red), urine mQTLs only (blue), and mQTLs in both biofluids (purple). Pie chart sizes are proportional to metabolite count. Metabolites without an AA mQTL were omitted. In total, 735 metabolites were measured in both biofluids, 639 in urine only and 581 in serum only. AA sample sizes are 818 for serum and 794 for urine. **(b)** KEGG pathway enrichment of prioritized genes significant in at least one biofluid. Pathways are grouped by significance pattern -serum-only (left), significant in both biofluids (center), and urine-only (right) -and sorted by fold enrichment ascending within each group. Color intensity reflects fold enrichment (**Methods**). An asterisk marks Bonferroni-corrected significance within each stratum.

To test whether this shift reflects power, we repeated the EA serum mGWAS in the 797 EA participants with paired urine measurements (**Supplementary Tables 6-7**; **Methods**). At matched sample size, the serum-specific proportion fell from 76% to 35%, urine-specific rose from 12% to 45%, and shared rose from 13% to 21%; the serum:urine mQTL ratio fell from 4.5:1 to 0.8:1, i.e. urine yielded more mQTLs than serum at equal N (**Extended Data Fig. 4**, **Supplementary Table 8**). All 115 power-matched mQTLs were recovered within 1 Mb of the original full-sample serum signals (median distance 4.4 kb), supporting that down-sampling reduces power without introducing spurious loci. These results indicate that the apparent predominance of serum-specific signals in the EA stratum reflects statistical power rather than biofluid-specific biology, and that at equal sample size urine is at least as informative as serum.

### The catalog replicates in independent cohorts and extends previous mGWAS

To place the catalog in the context of published literature, we compared all 969 mQTLs against eleven published mGWAS: nine Metabolon-based studies (Schlosser *et al.* ^14^, Chen *et al.*^6^, Long *et al.*^4^, Yin *et al.*^20^, Hysi *et al.*^26^, Lotta *et al.*^27^, Surendran *et al.*^5^, Shin *et al.*^3^, Luo *et al.*^16^) and, as secondary-tier evidence, two studies on other platforms (Tambets *et al.*^2^, Nuclear Magnetic Resonance; Tahir *et al.*^18^, untargeted Mass Spectrometry) (**Supplementary Table 9; Methods**).

Aligning the comparison to platform-matched studies of the same biofluid and ancestry, which could have detected a given signal, 289 of 969 mQTLs (30%) are not reported before, ranging from 9% in EA serum, the stratum most densely covered by prior work, to 30% in EA urine, 78% in AA serum and 100% in AA urine, where no prior same-platform study of that biofluid and ancestry exists (**Supplementary Table 9**). The 289 mQTLs not reported before include metabolites not measured in those studies, indels resolved by TOPMed-based imputation, and putative ancestry differences. Conservatively counting a report in any biofluid, any ancestry and any platform as prior knowledge, 88 of 969 mQTLs (9%) remain unreported.

Recovery was markedly higher within than across biofluids: ARIC urine mQTLs are recovered by other urine studies in 85% of cases versus 53% by serum/plasma studies, and ARIC serum mQTLs by other serum/plasma studies in 91% of cases versus 31% by urine studies, direct evidence for the biofluid complementarity that motivates this study.

As a targeted replication, we evaluated concordance with GCKD, the only other mGWAS with a paired blood-and-urine metabolomics design (**Supplementary Table 10; Methods**).^14^ In GCKD, 546 of 722 serum mQTLs (10 via proxy) and 181 of 247 urine mQTLs (8 via proxy) were testable (available variant and metabolite). Of these, 517 (95%) and 179 (99%) replicated with concordant direction after Bonferroni correction for the number of tested pairs. Effect estimates were highly concordant between studies (Pearson correlation coefficient = 0.83– 0.92; **Extended Data Fig. 5**).

Because neither GCKD nor any published resource covers urine metabolomics in African American participants, we obtained de-novo replication in the Chronic Renal Insufficiency Cohort^28,29^ (CRIC; N = 1,323 African American participants with urine metabolomics and genotyping; **Methods**). All 77 AA urine index variants were available at high imputation quality, and 102 of the 109 AA urine mQTLs were testable (seven of 101 metabolites were not measured in CRIC). All 102 (100%) showed a concordant direction of effect with ARIC and 93 (91%) replicated (Bonferroni correction for the 102 tests; **Supplementary Table 11**). Because CRIC is a chronic kidney disease cohort (mean eGFR 40.3 mL/min/1.73 m²), we pre-specified an eGFR-adjusted sensitivity model, which gave essentially unchanged results (95 of 102 Bonferroni-significant) (**Supplementary Table 11**).

Associations were also robust to adjustment for correlated metabolites. Re-analyzing the top locus at each of the 247 prioritized genes in each ancestry-by-biofluid stratum (366 gene-by-stratum combinations) with a method for covariate selection in multiphenotype studies (CMS; **Methods; Supplementary Table 12**) increased power at nearly every locus (median equivalent sample-size increase (SSincr) 1.48-fold), with effect estimates highly concordant with the marginal model (effect-size correlation r = 0.998; direction unchanged for all 366; median absolute relative change 4.3%). The largest gains were at *PON1*, a xenobiotic-and lipid-metabolizing enzyme, in both ancestries tested (serum SSincr 3.48-fold in EA and 3.37-fold in AA, for different target metabolites in each), consistent with broader metabolic co-regulation among its substrates (**Supplementary Table 12**).

### Prioritized genes point to hepatic and renal enzymes and transporters

For each mQTL, we prioritized a single candidate causal gene using a ProGeM-based framework integrating proximity, LD overlap, Variant Effect Predictor (VEP) consequences, cis-eQTL and cis-pQTL colocalization, and Polygenic Priority Score (PoPS) (**Methods**). Across all 247 unique prioritized genes, enzyme-class and transporter annotation showed a broad functional spread dominated by hydrolases, oxidoreductases and transferases, with transporters comprising 11.3% (**Extended Data Fig. 6**). The 220 and 81 genes associated with mQTLs in serum and urine were enriched for genes highly expressed in liver and kidney among other key organs involved in absorption, distribution, metabolism, and excretion (ADME; **Supplementary Tables 13-14**). For example, more than half of the genes were among the top 10% most specifically expressed genes in liver (ranked by tissue-specificity t-statistic across GTEx v8 tissues) with kidney cortex and medulla being the next most pronounced tissues.

Comparing KEGG pathway enrichment between serum and urine mQTL gene sets revealed both shared and biofluid-specific signals (**Supplementary Table 15**, **Fig. 3b**). Eight pathways were significantly enriched in both biofluids after Bonferroni correction, including drug and xenobiotic metabolism, steroid hormone biosynthesis, carbon metabolism, and amino acid catabolism. Among these, glyoxylate and dicarboxylate metabolism showed the highest urine fold enrichment (FE = 23.0), driven by *AGXT2*, *HAO2*, *AFMID*, and *GLDC*, with substantially stronger enrichment in urine than serum (urine/serum FE ratio = 2.5). Alanine, aspartate and glutamate metabolism was similarly enriched in both biofluids (serum FE = 12.6, urine FE = 19.3), with *AGXT2*, *GOT2*, and *CPS1* among the overlapping genes. Twenty pathways were significant exclusively in serum, predominantly involving xenobiotic processing, lipid and fatty acid metabolism, and bile secretion, consistent with the hepatic expression profiles of the underlying enzyme genes. Two pathways reached significance exclusively in urine, glutathione metabolism and butanoate metabolism, both pointing to renal tubular metabolic activity. Beta-alanine metabolism was enriched in serum but did not reach Bonferroni significance in urine despite high FE (FE = 17.25); notably, the sub-pathway enrichment of AA-specific mQTL-associated metabolites for pyrimidine metabolism (uracil-containing metabolites; *P* < 0.05/125 sub-pathways; **Supplementary Table 16**) directly complements this finding, as beta-alanine is a direct downstream product of pyrimidine (uracil) catabolism.

Repeating this enrichment analysis on the power-matched EA serum data (N=797) recovered the core biological signal at reduced power: 4 of 6 significant GTEx tissues were retained (liver, kidney cortex, small intestine and kidney medulla), 18 of 25 testable KEGG pathways were retained (3 pathways became untestable), with xenobiotic/drug-metabolism, bile secretion, and steroid-hormone-biosynthesis themes remaining robust while most individual amino-acid catabolism pathways (tryptophan, phenylalanine, valine/leucine/isoleucine, beta-alanine) lost significance. Sub-pathway resolution was far more power-sensitive, with only 1 of 9 full-sample-significant sub-pathways (Fatty Acid, Dicarboxylate) remaining significant. Broad pathway-level conclusions are therefore largely robust to the EA serum-urine power imbalance, while finer-grained pathway claims should be interpreted with this caveat (**Supplementary Tables 17–19**).

### Integrating European American and African American Ancestry Data to Enhance Genetic Discovery

Effect sizes replicated well between EA and AA in cross-lookups of the mQTLs (**Fig. 2c,d**). Power to detect these shared effects, however, differed substantially between ancestries because of differences in allele frequency. Of the 7.0 million genetic variants shared between the EA and AA analysis, 637,000 shared variants differ in MAF by ≥0.2 between ancestries (e.g., MAF_EA_ of 5% and MAF_AA_ of 25% or vice versa). For a sample size of 800 and effect size of 0.4, increasing the MAF from 0.05 to 0.25 would increase the power to detect this effect size from 2.5% to 93% (37-fold gain in power; **Fig. 4a**). In addition, there were 1.5 million and 7.9 million variants unique to the EA and AA analysis, respectively (**Fig. 4a**). At index variants of EA-discovered mQTLs, minor allele frequencies were systematically higher in AA than in EA (serum: *P* = 0.006; urine: *P* = 2.2 × 10⁻⁴). The same pattern held for index variants of AA-discovered mQTLs (serum: *P* = 7.7×10⁻⁵; urine: *P* = 1.1×10⁻⁹; **Fig. 4c**). In other words, mQTLs identified in either ancestry tended to sit at variants that are more common in AA, consistent with statistical power rather than ancestry-specific biology driving ancestry-specificity.

**Figure 4:**
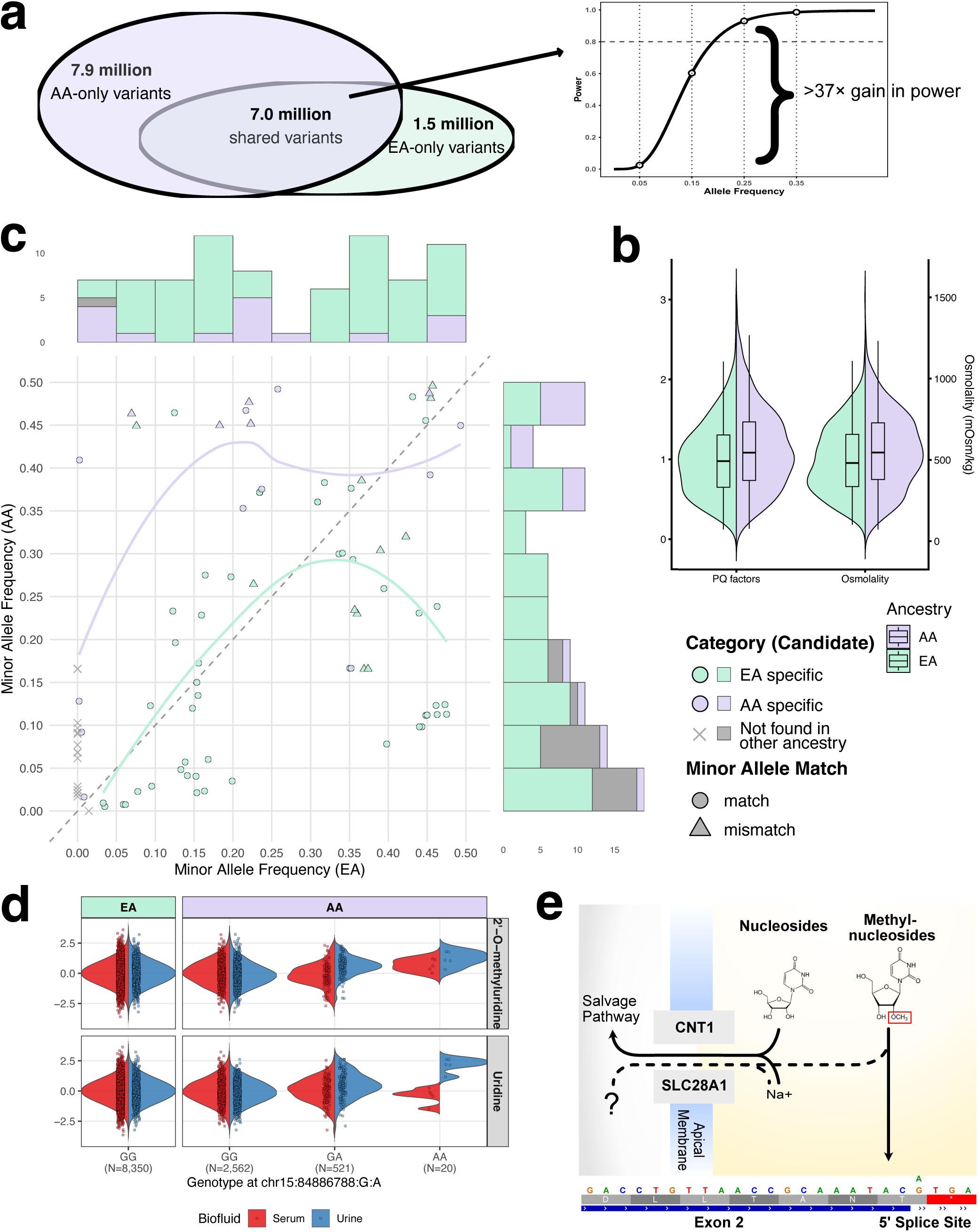
Advantages of multi-ancestry metabolite quantitative trait locus (mQTL) discovery. **(a)** Common variant (MAF>1%) availability across ancestry groups after TOPMed imputation: 7.9 million variants observed only in African Americans (AA), 1.5 million only in European Americans (EA), and 7.0 million shared between both groups. Inset: a variant is easier to detect in the ancestry where it is more common illustrated for a fixed effect and sample size (**Methods**). **(b)** Paired box-and-violin plots comparing the PQ factors (left) and osmolality provided by Metabolon (right) in EA versus AA participants. **(c)** Minor allele frequencies in EA (x-axis) versus AA (y-axis) for shared index variants in urine, with marginal histograms displaying the frequency distributions for candidate ancestry-specific variants (red: AA-specific; blue: EA-specific). Circles mark variants where the same allele is the minor allele in both ancestries; triangles mark variants where this differs. Trend lines (LOESS) are shown separately for EA-associated and AA-associated mQTLs. Gray crosses mark variants absent from the opposing ancestry. **(d)** Distribution of 2’-O-methyluridine and uridine levels by genotype at chr15:84886788:G:A in EA (left) and AA (right) participants. Split violins (serum: red, left half; urine: blue, right half) show density per genotype class, with individual jittered points colored by biofluid. The variant is monomorphic in EA (GG only), while AA carriers show three genotype classes (GG, GA, AA). **(e)** Functional example: a splice-region variant in *SLC28A1*, encoding a sodium-coupled nucleoside transporter expressed at the apical membrane of renal proximal tubule cells. The variant affects the 5ʹ splice site of exon 2, potentially disrupting transporter function and altering nucleoside salvage pathway activity.

A notable example of findings in which MAFs were greater among the EA population was a clustering of urine mQTLs with MAF_EA_>0.4 and MAF_AA_<0.15 annotated to *SLC17A1-3*(**Fig. 4c**) and attributed to the power-related candidate-ancestry-specific mQTL. These mQTLs represent genetic regulation of several glutamine derivatives at a locus well-known for its pathogen-driven selection.^30^ Moreover, power to detect urinary mQTLs might also be influenced by the urine concentration. We observed higher osmolality and PQ factors for the AA sample (*P*_PQ-factors_ = 3.3 × 10^-7^; **Fig. 4b**; Methods).

Across both biofluids, 632 mQTLs were candidate ancestry-specific (590 EA-specific, 42 AA-specific; **Supplementary Tables 3-4**), including 95 of the 247 urine mQTLs (62 EA-specific, 33 AA-specific) despite comparable sample sizes of the 159 implicated metabolites across ancestries (range sample size difference between ancestries = [-7.6%, 5.9%]). To evaluate whether these differences may be attributable to statistical effects of unequal variant availability or power, we applied a three-step classification based on variant availability, power in the other ancestry and effect-size heterogeneity (Methods; **Extended Data Fig. 2**). Of the 632 candidates, 609 (96%) were attributed to variant unavailability or insufficient power in the other ancestry. Only 23 (4%) showed significant heterogeneity irrespective of sample size and MAF and are termed ancestry-specific (**Supplementary Table 20**); three of these (*UNC93A*, *LCOR* and *SLC36A2*) are also fine-mapped ancestry-specific signals in MESuSiE.

### An African American-enriched splice variant in SLC28A1 implicates renal nucleoside reabsorption

In AA, we identified an mQTL with the index variant chr15:84886788 (G>A; β = 0.64, *P* = 2.82 × 10⁻¹³, N = 772, MAF=0.09) strongly associated with urinary 2’-O-methyluridine levels as well as urinary uridine (β = 0.62, *P* = 9.72 × 10⁻¹³, **Fig. 4d**), with negligible frequency in EA (gnomAD v4.1: AFR MAF = 8.5%, EUR MAF ≈ 0%, **Fig. 4d**), so that this candidate ancestry-specific mQTL falls into the AF-driven class above, while MESuSiE assigns it an AA-specific credible set (fine-mapped ancestry-specific). This variant was independently confirmed as a liver-specific splice quantitative trait locus (splice-QTL) in the GTEx Project (P = 1.2 × 10⁻²⁸). The variant disrupts the canonical 5’ splice donor motif immediately following exon 2, predicted to result in exon 2 skipping and consequent alteration of the 5’ untranslated region.^31^ Together, this genetic and functional evidence indicates that the *SLC28A1* locus variant results in reduced expression of the concentrative nucleoside transporter CNT1 encoded by the gene (**Fig. 4e**). Consistent with the canonical CNT1 substrate spectrum, the effect allele was additionally associated with reduced serum cytidine in African American participants, paralleling the pyrimidine nucleoside excretion phenotype observed in *Slc28a1⁻/⁻* mice^32^ and in two siblings with biallelic loss-of-function *SLC28A1* variants (p.Ser546Pro), who showed elevated urinary uridine, cytidine, deoxycytidine, and 5-methylcytidine.^33^ The concordant increase in urinary uridine and 2’-O-methyluridine in our study extends these observations, as methylated pyrimidine derivatives including 5’-methyldeoxycytidine, 3-methylcytidine, and N4-acetylcytidine were likewise elevated in the urine of CNT1-deficient mice^32^.

### Allele-frequency differences rather than selection underlie most ancestry differentiation

Even in serum, where the EA sample was 4.4-fold larger, we identified nine candidate AA-specific mQTLs (**Supplementary Table 4, Extended Data Fig. 7**). Six of these index variants were not available in EA, including those of three fatty acid dicarboxylates associated with *ADH1B* (MAF 22% in AA). Of the three available in EA, two showed large MAF differences: 3beta,7alpha-dihydroxy-5-cholestenoate at *HSD3B7* (31% in AA vs 1% in EA) and 3-methoxytyrosine at *DDC* (15% in AA vs 1% in EA). The remaining variant, associated with N6-methyllysine at *LCOR*, had a comparable MAF in both ancestries (16% in AA vs 19% in EA).

A notable example illustrating the impact of ancestry differences in allele frequency on mQTL discovery is the missense variant rs37369 (chr5:35037010:C:T; NM_031900.4:c.418G>A, p.Val140Ile) in *AGXT2*, encoding alanine-glyoxylate aminotransferase 2. This variant was associated with 3-aminoisobutyrate levels across both biofluids in EA, where both associations were classified as shared between ancestries because of an AA mQTL within the 1 Mb window. In total, 3-aminoisobutyrate had three serum mQTLs (1 EA-AA shared, 1 candidate EA-specific) and two urine mQTLs (1 AA-EA shared) of which only the candidate EA-specific was not annotated to *AGXT2* but rather to *SLC6A13*. The effect allele (T, derived allele) frequency of rs37369 differs markedly between populations, at approximately 8% in EA and 55% in AA, with substantial global variation reported across diverse populations. To our knowledge, no published genome-wide scans have reported evidence of positive selection at this locus (Fst = 0.062, 76th genome-wide percentile, empirical *P* = 0.24), and the observed frequency difference most likely reflects drift rather than adaptive divergence. This frequency disparity has direct consequences for statistical power: while the variant reached genome-wide significance for 3-aminoisobutyrate in EA in both biofluids, the weaker serum effect failed to reach significance in AA, and the urine association was only marginally genome-wide significant (*P* ≈ 1 × 10⁻⁹). The variant replicated in GCKD, where the effect allele frequency (∼8%) closely mirrors that of ARIC EA, consistent with the European ancestry of that cohort. In the locus-level classification, the 3-aminoisobutyrate mQTLs at *AGXT2* are shared between ancestries in both biofluids (**Supplementary Tables 3-4**), so the frequency difference modulates power at a shared locus rather than marking metabolite handling that works differently between ancestries.

A contrasting case is the *UNC93A* locus (chr6:167294271:C:T), one of the 23 loci that remain ancestry-specific after the classification above and also a fine-mapped ancestry-specific signal. *UNC93A* was recently nominated, by a machine-learning screen of orphan solute carriers, as a candidate transporter for acetylglucosamine; cells overexpressing *UNC93A* showed markedly increased cellular acetylglucosamine uptake, functionally supporting this candidacy, and its genetic association with circulating acetylglucosamine had previously been reported in European-ancestry cohorts.^34^ In our data, the effect allele is common in both ancestries (AA: 17.6%; EA: 10.7%) and shows a strong, GCKD-replicated association with serum *N*-acetylglucosamine/*N*-acetylgalactosamine in EA (β = 0.455, *P* = 4.84 × 10⁻³⁵), yet no association at all in AA (β = −0.008, *P* = 0.89, adequately powered at *N* = 818). Finemapping resolves the locus to a single variant (PIP > 0.999), making differential linkage disequilibrium an unlikely explanation for this heterogeneity; a genuine gene-by-ancestry or gene-by-environment interaction is more plausible, though distinguishing between these would require direct functional follow-up. Dietary or microbiome-driven differences in circulating acetylglucosamine are one possible explanation that warrants further investigation. *UNC93A* thus illustrates how population-specific genetic regulation can be layered onto a transporter whose basic substrate biology has only just been established.

We next asked whether the frequency differentials extend across the mQTL catalog, using Fst between reference populations as a measure of allele-frequency differentiation (**Methods**). Fst was available for 609 of 969 index variants (63%, **Supplementary Tables 3-4**); for each, we report its percentile in the genome-wide Fst distribution, i.e., the share of all variants that are *less* differentiated between CEU and YRI. Differentiation is only modestly elevated overall (mean percentile 57.9; 9.0% of variants in the top 5% of the genome-wide distribution, against 5% expected). Index variants of mQTLs identified in African American participants sit at a higher mean percentile than those identified in European American participants (66.1 vs. 55.9), driven mainly by signals shared across ancestries (69.2) rather than those specific to African American participants (54.7). Because allele-frequency differentiation can arise from drift as well as from selection, these data indicate at most a weak, nonpervasive contribution of population-differentiating selection to the frequency differentials described above.

### Multi-ancestry finemapping resolves candidate causal variants and genes

We utilized MESuSiE^35^ to finemap 992 independent signals underlying 627 unique non-overlapping region-metabolite combinations implicated by serum mQTLs as well as 238 independent signals underlying 167 regions in urine (**Supplementary Tables 21-24, Supplementary Data 1**). The multi-ancestry analysis improved the ability to detect independent signals when compared to ancestry-specific finemapping using the same model (SuSiE, **Methods**; **Fig. 5a, Extended Data Fig. 8**). We were able to narrow down many of the EA credible sets by incorporation of evidence from the AA sample -even for the serum analysis with the larger EA sample size (**Fig. 5c**). Multi-ancestry finemapping improved credible set resolution compared to single-ancestry SuSiE (median set size: 2 vs. ≥6), with AA-specific regions showing the smallest sets, likely reflecting shorter linkage disequilibrium blocks (mean signals per region: 1.95 vs. ≤1.46 for all other ancestry-method pairs).

**Figure 5:**
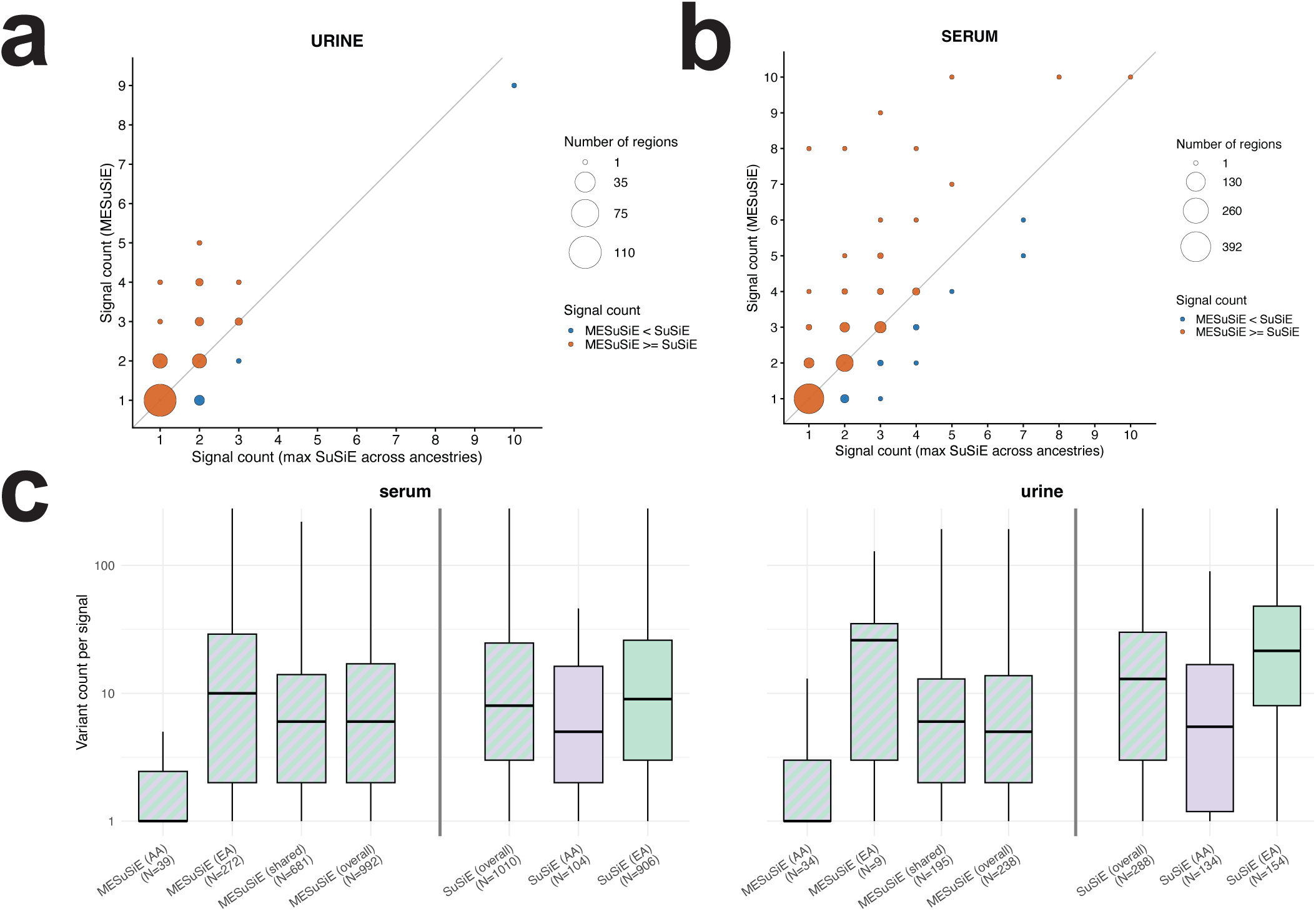
Multi-ancestry statistical finemapping of mQTL regions. The number of independent signals multi-ancestry fine-mapping (MESuSiE) finds per region, compared to the best single-ancestry result (SuSiE), for urine **(a)** and serum **(b)**. Circle size indicates the number of regions; most regions lie on or above the diagonal, meaning multi-ancestry fine-mapping finds at least as many signals as single-ancestry analysis alone. **(c)** Credible set sizes (number of variants per signal, log scale) for MESuSiE and SuSiE across serum (left) and urine (right). MESuSiE credible sets are stratified by ancestry assignment (AA-specific, EA-specific, or shared), with overall totals shown for both methods. Color indicates whether the source data originated from EA, AA, or both ancestries. Smaller credible sets reflect greater resolution in localizing causal variants.

Among the 759 unique finemapped variants with high posterior inclusion probability (PIP) or part of small credible sets in either biofluid (credible set size ≤5 or PIP ≥0.5; **Supplementary Tables 22 and 24**), 713 were SNVs and 46 were indels. Compared to a random background set of 10,000 common variants, finemapped variants showed a markedly different consequence profile (**Fig. 6a**). VEP annotated the majority as MODIFIER impact (87.2%), consistent with their predominantly non-coding genomic context. Notably, 9.4% of finemapped SNVs were classified as missense, and 0.8% of all variants carried HIGH impact consequences, including splice donor variants. The proportion of HIGH impact variants was highest among indels (4.3%), followed by SNVs (0.4%), with a background rate of 0.0%, highlighting the potential of multi-ancestry-based statistical finemapping to prioritize functionally consequential variants. To complement categorical annotation with quantitative predictions of regulatory impact, we scored all finemapped variants using AlphaGenome,^36^ a deep learning model that predicts molecular effects across chromatin accessibility, transcription factor binding, histone modification, and gene expression tracks in hundreds of cell types (**Supplementary Tables 22 and 24**). As with other in-silico predictions used for gene and variant prioritization throughout this study, AlphaGenome scores are hypothesis-generating and were not experimentally validated here. Of the 759 variants, 746 (98.3%) showed at least one high-confidence predicted effect (|quantile score| > 0.99; **Methods**; **Supplementary Note 1**). A total of 251 variants (33.1%) showed predicted disruption of at least one transcription factor binding site in any tissue. Several established regulators of hepatic and renal gene expression were among the most frequently disrupted transcription factors: SP1 (53 variants, 21.1% of TF-hit variants), CEBPB (43, 17.1%), RXRA and FOXA1 (35 each, 13.9%), FOXA2 (32, 12.7%), and HNF4A (25, 10.0%). Of these, HNF4A, FOXA1, FOXA2, and RXRA are liver-enriched transcription factors that cooperatively regulate metabolic gene programs in liver and kidney^37,38^, while SP1 and CEBPB are broadly expressed transcription factors with known roles in hepatic gene regulation^39,40^. When restricting to metabolically relevant tissues (**Methods**), kidney was the most frequent top organ (228 of 687 annotated variants, 33.2%), followed by liver (176, 25.6%), consistent with the renal and hepatic origin of many circulating and urinary metabolites.

**Figure 6:**
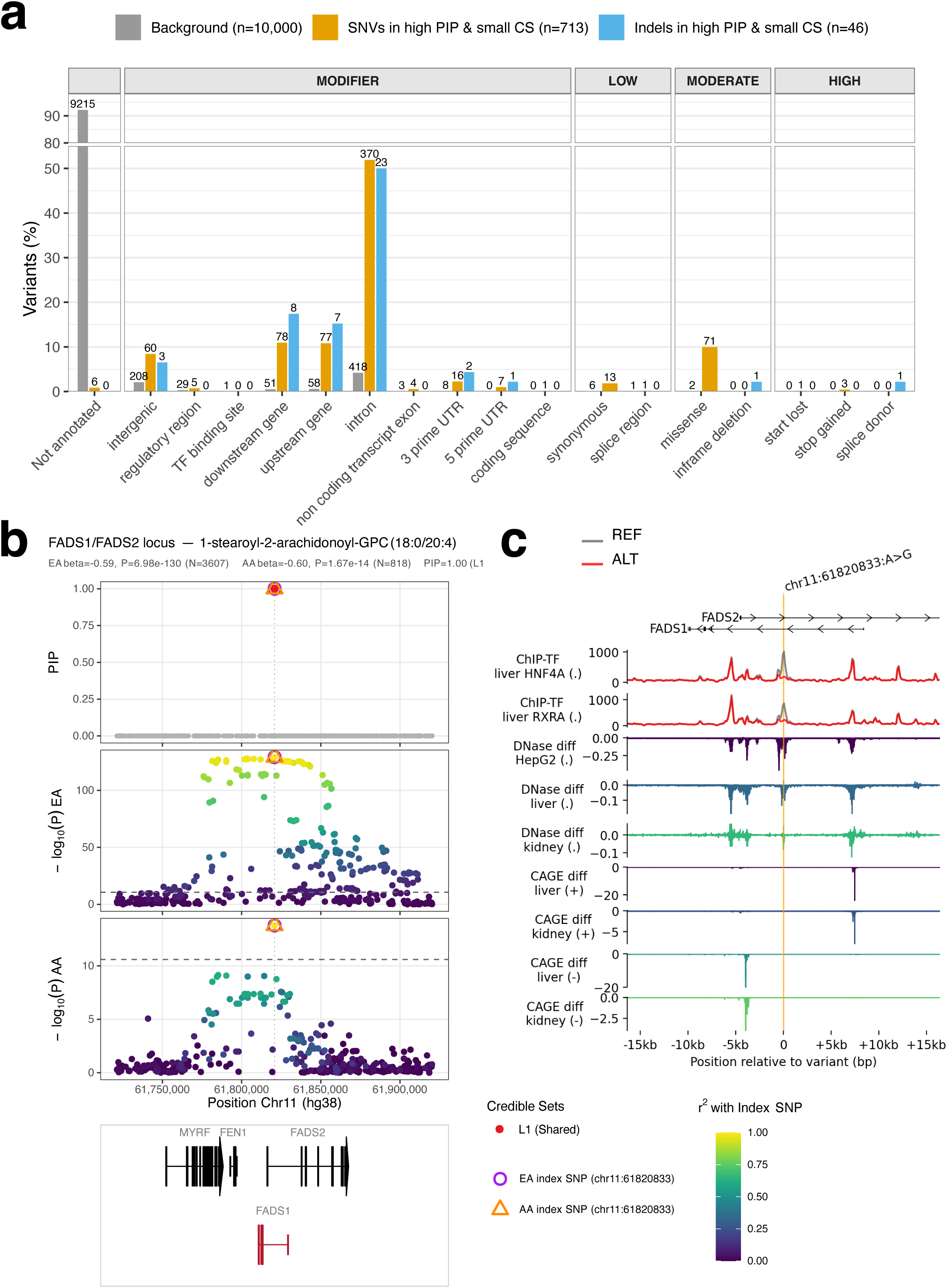
Functional and regulatory characterization of finemapped variants. **(a)** Distribution of VEP-predicted most severe consequences among finemapped mQTL variants. Grouped bar chart showing the percentage of variants annotated with each Variant Effect Predictor (VEP) consequence type (most severe), stratified by variant class: background variants (gray; random subset of common variants, n=10,000), and finemapped SNVs (orange, n=713) and indels (blue, n=46) meeting the high-PIP criterion (**Methods**). Consequences are ordered by increasing VEP-defined severity and grouped by VEP IMPACT category (MODIFIER, LOW, MODERATE, HIGH; facet labels above). Bars for missense and synonymous variants are omitted for the indel group, as these consequence types apply exclusively to single nucleotide substitutions. The y-axis is broken between 55% and 80% to accommodate the high proportion of unannotated background variants. **(b)** Regional Association Plot of the *FADS1/FADS2* locus. From top to bottom: (1) Posterior Inclusion Probabilities (PIPs) from MeSuSiE multi-ancestry fine-mapping. Variants belonging to credible sets are colored by credible set membership (L1, L2, etc.); remaining variants are shown in gray. The EA index variant is indicated by a purple circle and the AA index variant by an orange triangle. (2, 3) Regional association in the European-and African American-ancestry populations, respectively, showing −log10(P) derived from MeSuSiE Z-scores. Points are colored by r² with the corresponding ancestry’s index variant (gray: no LD information). (4) Gene models for the locus. **(c)** AlphaGenome-predicted variant effects at the *FADS1/FADS2* locus (chr11:61820833:A>G). Top: gene annotation showing the head-to-head arrangement of *FADS2* (positive strand) and *FADS1* (negative strand). Panels show the predicted effect of the alternate allele: reduced binding of two liver transcription factors (HNF4A, RXRA); reduced chromatin accessibility in HepG2 cells, liver, and kidney; and differences in transcription initiation on both DNA strands. X-axis shows position in base pairs relative to the variant.

Among the 251 unique high-PIP variants with predicted transcription factor binding disruption, 25 (10.0%) were predicted to affect HNF4A binding in primary liver tissue or HepG2. HNF4A is of particular biological interest given its established role as a master regulator of hepatic lipid and fatty acid metabolism. One such variant, chr11:61820833:A:G (rs174564), annotated by VEP as an upstream gene variant of *FADS1* on chromosome 11, disrupts a predicted HNF4A binding site in liver and appeared in credible sets for 17 serum metabolites spanning phosphatidylcholines, plasmalogens, diacylglycerols, and free arachidonate -all substrates or products of the *FADS1/2* desaturation pathway. For ten of these metabolites, the variant was the sole member of the credible set (PIP > 0.96, **Supplementary Table 22**), making it one of the most confidently finemapped multi-metabolite variants in our study. Incorporated annotation resources indicate an involvement of *FADS1*, *FADS2*, and *FEN1* in the region with multiple colocalization signals with gene expression of all three genes. For EA mQTLs associations were mapped to *FEN1* for 9 metabolites and *FADS1* for 7, while the AA gene assignment, available for 4 metabolites (3 shared, 1 AA-specific), consistently pointed to *FADS1*, including one AA-specific singleton credible set for 1-stearoyl-2-arachidonoyl-GPC (18:0/20:4), a phosphatidylcholine containing arachidonic acid (PIP = 1.00). This locus illustrates directly how multi-ancestry finemapping improves resolution: for this metabolite, the EA association is exceptionally strong (*P* = 6.98 × 10⁻¹³⁰) but falls within a broad region of high linkage disequilibrium, with several correlated variants reaching comparably high significance and no single candidate resolved when EA is considered alone. The AA association, while considerably weaker (*P* = 1.67 × 10⁻¹⁴), shows a more localized pattern around the same variant; jointly modeling both ancestries with MESuSiE collapses the credible set to this single variant (PIP > 0.999), despite AA’s lower power (**Fig. 6b**).

AlphaGenome predicted that the alternate allele disrupts binding of multiple hepatic transcription factors at this site, most strongly HNF4A and RXRA in liver (**Fig. 6c, Supplementary Table 22**). Experimental support for HNF4A binding at this site is provided by ChIP-seq data curated in UniBind^41,42^, which identifies robust, quality-controlled HNF4A and RXRA binding sites overlapping rs174564 in primary liver tissue, with additional HNF4A occupancy observed in HepG2 cells. AlphaGenome predicted reduced DNase accessibility across liver, kidney, and HepG2, consistent with chromatin closure at the variant position. Notably, the CAGE transcription initiation tracks revealed a strand-dependent effect: on the positive strand (corresponding to *FADS2*), predicted CAGE signal was reduced in both liver and kidney, whereas on the negative strand (corresponding to *FADS1*), the reduction was concentrated at a distinct upstream promoter element (**Fig. 6c**). This strand-specific CAGE pattern suggests that the variant may simultaneously affect the transcriptional regulation of *FADS1* and *FADS2* on their respective strands, consistent with the known divergent transcription of this head-to-head gene pair and the established role of HNF4A in regulating hepatic fatty acid desaturase expression. The predictions also suggest additional regulatory effects at the *FEN1* transcription start site (full 1 Mb prediction window in **Extended Data Fig. 9**). This locus also showed the broadest disease colocalization in our catalog (see below).

The *ALDH7A1* locus on chromosome 5 illustrates how combining multi-ancestry finemapping with deep learning-based functional prediction can strengthen the case for follow-up of individual mQTL variants. Two mQTL regions were identified for serum levels of 6-oxopiperidine-2-carboxylate, a key intermediate in lysine catabolism, which accumulates in the plasma and urine of patients with *ALDH7A1* deficiency: a shared signal at the *OPLAH* locus and an EA-specific signal at *ALDH7A1* on chromosome 5, encoding aldehyde dehydrogenase 7 family member A1. At the *ALDH7A1* locus, the index variant was not represented in the 1000 Genomes CEU/YRI reference panel and therefore lacks an Fst estimate, consistent with its classification as a candidate ancestry-specific signal driven by variant unavailability in EA. The EA association was strong (β = 0.54, *P* = 6.45 x 10^-19^, N = 3,595) whereas no significant signal was detected in AA (β = 0.28, *P* = 0.31, N = 811), consistent with a pronounced allele frequency difference (EAF_EA_ = 0.04 vs. EAF_AA_ = 0.01). Finemapping identified a single credible set of five variants with a cumulative coverage probability of 0.97. The top variant chr5:126595258:C:T carried a PIP of 0.52. This variant is also the EA index variant for the locus (**Extended Data Fig 10a**). Finemapping classified the credible set as EA-specific, underscoring how the multi-ancestry framework helps delineate ancestry-dependent genetic architecture even when one stratum contributes minimal information for finemapping above LD patterns. VEP annotated the variant as an upstream gene variant (CADD PHRED score 18.8). The variant is classified as benign/likely benign in ClinVar with respect to Mendelian disease, consistent with its role as a common regulatory variant rather than a disease-causing allele. AlphaGenome predicted tissue-differential regulatory disruption across multiple functional modalities. In the ChIP-TF output, the alternate allele was associated with reduced predicted binding of RNA Polymerase II (POLR2A) across 42 biosamples, as well as diminished binding of liver-relevant transcription factors including SP1, FOXA1, FOXA2, and RXRA (**Supplementary Table 22**). Notably, the predicted effect on the H3K4me3 active promoter mark was tissue-dependent: in K562, the variant was associated with a pronounced loss of H3K4me3 at the *ALDH7A1* promoter, whereas in liver and kidney -the primary tissues of *ALDH7A1* expression and lysine catabolism -H3K4me3 was predicted to increase (**Extended Data Fig. 10b**). By contrast, AlphaGenome predicted uniformly reduced DNase-seq accessibility at the variant site across all three tissues, suggesting that the variant alters chromatin accessibility broadly but exerts tissue-specific effects on promoter histone modifications. The H3K4me3 gain in liver and kidney suggests that the variant may enhance promoter activity at *ALDH7A1* specifically in tissues central to lysine catabolism, consistent with the serum-specific mQTL association observed for its downstream product 6-oxopiperidine-2-carboxylate. Taken together, the convergence of a moderate-PIP finemapping signal with consistent, tissue-specific epigenomic disruption predicted by AlphaGenome supports chr5:126595258:C:T (*ALDH7A1*:c.-60G>T, a 5 prime UTR variant) as a high-priority candidate for experimental follow-up to understand the metabolic block in lysine catabolism, and illustrates how deep learning-based variant effect prediction can add functional confidence to statistically ambiguous finemapping results.

### Colocalization links the catalog to cardiometabolic, kidney and liver disease

To link our mQTL catalog to diseases and clinical outcomes, we conducted colocalization analysis to test whether each locus shares a causal variant with 3,037 trait-ancestry outcomes from the Million Veteran Program (MVP) phenome-wide GWAS (**Methods**).^17^ At a posterior probability of a shared causal variant (PP.H4) > 0.8, we detected colocalizations for 521 metabolite-stratum pairs (381 unique metabolites) across 125 genes and 217 MVP trait families spanning 12 disease categories (**Supplementary Table 25**). Quantitative traits dominated, led by serum triglycerides (182 metabolite-stratum pairs, 28 genes) and eGFR (181 metabolite-stratum pairs, 23 genes). Among binary outcomes, the strongest metabolite-level signals were gout (70 metabolite-stratum pairs, 12 genes), hyperlipidemia (73, 11), chronic kidney disease (74, 4), liver-function abnormalities (63, 6) and hypercholesterolemia (57, 14). The implicated diseases matched the known physiology of the prioritized genes. Genes involved in lipid transport and metabolism colocalized with triglycerides, cholesterol and hyperlipidemia (e.g. *ALDH1A2*, *UGT1A1*); kidney-expressed transporters and enzymes with creatinine, eGFR and chronic kidney disease (e.g. *NAT8*); and urate-handling loci with gout (e.g. *ABCG2*, *SLC22A5*, *GCKR*). The most broadly connected genes were *FADS1*, a fatty-acid desaturase (31 metabolite-stratum pairs, 63 trait families spanning lipid, cardiovascular and inflammatory traits), whose signal multi-ancestry finemapping resolved to a single candidate variant (see above); *SLCO1B1*, a hepatic transporter of bile acids and statins (45 metabolite-stratum pairs, 20 trait families including liver and lipid phenotypes); and *CPS1*, the rate-limiting enzyme of the urea cycle (26 metabolite-stratum pairs, 13 families centered on kidney and nitrogen-handling traits). That the diseases recovered for each gene consistently match its established function gives independent support to our gene prioritization, beyond metabolite-level evidence alone.

A locus at *ENPEP* (aminopeptidase A, chr4:110.42–110.57 Mb), which cleaves angiotensin II in the renin-angiotensin system, colocalized with fibrinopeptide A degradation fragments (thrombin-cleavage products of fibrinogen, distinct from fibrinogen itself) and with blood pressure, hypertension and serum potassium traits (PP.H4 up to 1.00). The lead variant, a rare stop-gain allele in *ENPEP* (rs33966350), has independently been shown to reduce kidney *ENPEP* expression and raise hypertension risk,^43^ and an association between this same variant and fibrinopeptide A fragments has been reported in an Amish cohort^44^ (**Extended Data Fig. 11**).

We investigated this locus further with complementary medication-based observational analyses in ARIC (**Methods, Supplementary Note 2**, **Extended Data Fig 11**). These indicate that genetically or pharmacologically reduced ACE activity specifically alters fibrinopeptide fragment levels. ACE-inhibitor use (but not angiotensin-receptor-blocker use) reproduces this pattern (e.g. FPB(1-13), *P* = 2 × 10⁻²⁹⁶) and in a mechanistically coherent direction, while clinical coagulation markers (fibrinogen, APTT, von Willebrand factor), available only from earlier study visits, did not differ between later ACE-inhibitor users and untreated participants in the same cohort. A plausible explanation is that ACE continues to act on the fibrinopeptide substrate pool generated during clot formation within the serum sample tube, similar in principle to a synthetic-substrate *ACE* activity assay, so that *ACE* genotype and ACE-inhibitor status at the time of blood draw are reflected in these measurements alongside any physiological signal.

A second locus, at *SLC16A9* (MCT9, a renal carnitine/urate transporter, chr10:59.67–59.71 Mb), colocalized with serum acylcarnitines, urate-related traits (gout, PP.H4 up to 0.998) and coronary atherosclerosis (PP.H4 up to 0.997) simultaneously (**Extended Data Fig. 12**). A carnitine-mediated mechanism was biologically plausible a priori: MCT9 mediates carnitine efflux in a context-dependent manner,^45^ and altered cellular carnitine handling could affect mitochondrial fatty-acid oxidation and acylcarnitine accumulation, both previously linked to cardiac lipotoxicity. Mendelian randomization (**Methods, Supplementary Note 3**) supported a causal effect of urate on gout and, less robustly, on coronary artery disease (significant with the inverse-variance-weighted and weighted-median estimators but not with MR-Egger), mirroring previous Mendelian randomization analyses of urate and coronary heart disease.^46^ At this locus, the carnitine-and urate-transport signals were not statistically separable, so the coronary colocalization cannot be attributed to carnitine (see Discussion). A genome-wide gout instrument was similarly associated with coronary artery disease, but showed directional pleiotropy (MR-Egger intercept test), likely reflecting that a case-control gout diagnosis captures broader inflammatory/comorbidity-driven biology beyond urate handling, rather than a clean urate-specific signal.

## Discussion

Our multi-ancestry genome-wide association study of paired serum and urine metabolomes in the ARIC cohort represents, to our knowledge, the first systematic investigation combining ancestrally diverse populations with multi-biofluid metabolite profiling. We identified 969 mQTLs for 553 metabolites, replicated them in GCKD and, for urine mQTLs in the African American stratum, de novo in CRIC, and linked 521 metabolite-stratum pairs to clinical traits through colocalization with the MVP phenome. Three findings stand out. Serum and urine capture complementary genetic regulation that is not explained by sample size. Most ancestry-differentiated signals reflect allele frequency and power rather than ancestry-specific metabolite handling, yet frequency differences expose functional variants such as the *SLC28A1* splice variant. Multi-ancestry finemapping and colocalization together prioritize enzymes and transporters whose disease links match their physiology.

The contribution of this catalog does not rest on locus counts alone. Looking at biofluid-and ancestry-matched studies using Metabolon, 30% of our mQTLs are not reported before, rising to 100% in the urine African American stratum, where no comparable prior resource exists, and 9% remain unreported even when a report in any biofluid or ancestry or platform is counted as prior knowledge. What cannot be obtained by combining the existing literature is the design itself: paired serum and urine metabolomes measured in the same individuals, a combination only GCKD also provides, and only in a chronic-kidney-disease population, together with a substantial African American component in both biofluids, for which essentially no prior Metabolon urine resource exists.

The complementary nature of serum and urine metabolome profiling was a central finding of our study. In the African American cohort, where sample sizes were balanced across biofluids, we observed that 36% of metabolites with mQTLs had associations detectable only in serum, 39% only in urine, and 25% in both biofluids. This distribution aligns with proportions previously reported in the GCKD study, which demonstrated that urine metabolomics can reveal genetic associations invisible to studies focusing solely on circulating metabolites.^14^ The apparent serum predominance in European Americans disappeared at matched sample size, and other studies recovered our mQTLs far more often within than across biofluids (urine: 85% versus 53%; serum: 91% versus 31%), directly quantifying how much biofluid-specific effects a single-biofluid design would miss.

These differences follow from physiology. Circulating levels integrate hepatic synthesis, conjugation and uptake, whereas urinary levels additionally reflect glomerular filtration and tubular secretion and reabsorption. Consistent with this, mQTL genes were enriched for liver and kidney expression; serum-only pathways centred on xenobiotic processing, lipid and fatty acid metabolism and bile secretion; and glutathione and butanoate metabolism were enriched only in urine, while glyoxylate and dicarboxylate metabolism showed its strongest enrichment in urine, pointing to renal tubular metabolism. A variant in a tubular transporter can therefore leave a large urinary signal with a smaller change in blood, as at *SLC28A1*, where the effect allele raised urinary uridine and 2’-O-methyluridine but lowered serum cytidine. The AA-specific enrichment of uracil-containing pyrimidine metabolites, upstream of beta-alanine, points to further regulation that single-ancestry studies would miss.

A key motivation for our multi-ancestry design was to leverage the genetic diversity present in African American populations to improve finemapping precision. African American populations exhibit shorter linkage disequilibrium (LD) blocks and greater genetic diversity compared to European populations, features that have been shown to substantially improve the resolution of credible sets in finemapping analyses.^35^ The Million Veteran Program has demonstrated that multi-ancestry approaches can identify population-specific causal variants and narrow credible sets relative to European-only analyses.^17^ Consistent with these expectations, our multi-ancestry MESuSiE finemapping detected more independent signals than single-ancestry SuSiE, and the African American-specific signals yielded the most specific credible sets. Even in serum, where European American sample sizes were 4.4-fold larger, incorporating African American data improved credible set resolution, as at *FADS1*, where the joint model resolved a signal that European-ancestry data alone could not.

Ancestry-differentiated discovery was largely a matter of allele frequency. Of 632 candidate ancestry-specific mQTLs, only 23 (4%) showed heterogeneous effects once variant availability and power were accounted for, and index variants discovered in either ancestry were systematically more common in African Americans. Population differentiation (Fst) at index variants was only modestly elevated and higher at ancestry-shared than at candidate ancestry-specific loci among African American, arguing against widespread selection. Diverse cohorts therefore reveal biology that is shared across ancestries but detectable only where the relevant allele is common, as at *SLC28A1*, or that modulate power at shared loci such as *AGXT2*. Genuine heterogeneity, as at *UNC93A*, was rare and may reflect gene-by-environment interactions, for example through diet or the microbiome.

Among loci shared across ancestries, the *AGXT2*/rs37369 region exemplifies how multi-ancestry mQTL mapping can illuminate population-specific variants with clinical relevance. The variant rs37369 has previously been identified as the genetic basis of hyper-beta-aminoisobutyric aciduria in a previous urine metabolomics GWAS,^9^ and subsequent work has confirmed that *AGXT2* polymorphisms, including this variant, have considerable impact on both methylarginine and beta-aminoisobutyrate metabolism. Beyond metabolomics, the variant and the *AGXT2* locus have been implicated in blood pressure regulation,^47^ coronary artery disease risk, and chronic heart failure^48^, with several of these findings originating from East Asian populations where the effect allele frequency more closely resembles that observed in AA than in EA. The pleiotropic involvement of *AGXT2* in both metabolite handling and cardiorenal phenotypes, combined with the pronounced population-level allele frequency variation at this locus, underscores the importance of multi-ancestry study designs for comprehensive mQTL characterization.

The *SLC28A1* locus exemplifies the power of multi-ancestry approaches to identify functional variants with population-specific effects. We detected an African Amercian-enriched splice-disrupting variant associated with urinary 2’-O-methyluridine levels -a signal that would have been missed entirely in European-ancestry-only studies due to the near-absence of this allele in European populations. *SLC28A1* encodes the concentrative nucleoside transporter CNT1, a sodium-dependent pyrimidine nucleoside transporter highly expressed in kidney tubular epithelium.^49^ Our genetic findings provide human evidence for the potential role of *SLC28A1* in methylated nucleoside transport, validating and extending observations from Slc28a1-knockout mice.^32^ The conserved metabolic phenotype across species underlines its physiological importance.

The functional consequences of *SLC28A1* loss have been characterized in both human disease and mouse models. Biallelic loss-of-function mutations in *SLC28A1* cause uridine-cytidinuria, an inborn error of metabolism characterized by elevated urinary excretion of pyrimidine nucleosides.^33^ Supporting our findings, metabolomic profiling of *Slc28a1*-knockout mice demonstrated elevated urinary excretion of both canonical pyrimidines and methylated pyrimidine derivatives, suggesting that modified nucleosides derived from tRNA and rRNA degradation are also low-affinity substrates of this transporter.^32^ Our observation that the African American-specific splice variant was associated with both increased urinary 2’-O-methyluridine, urinary uridine and reduced serum cytidine levels is consistent with impaired tubular reabsorption of pyrimidine nucleosides. Whether methylated nucleosides are direct CNT1 substrates (consistent with CNT1’s established capacity to transport structurally modified nucleoside analogues) or accumulate secondarily to elevated parental nucleoside pools and downstream modification cannot be distinguished from association data alone. The supporting evidence from a liver-specific splice-QTL in GTEx^50^ strengthens the case for this variant’s functional impact.

This finding may have broader implications for pharmacogenomics, as CNT1 is involved in the disposition of nucleoside analog drugs, including gemcitabine and other antineoplastic and antiviral agents.^51^ Population-specific functional variants in drug transporters may contribute to inter-individual variability in therapeutic response and toxicity, highlighting the importance of including diverse populations in both basic science and translational research.

Our use of TOPMed-based imputation contributed importantly to our results.^52^ The TOPMed reference panel, with its substantial inclusion of African and Hispanic/Latino individuals, provides superior imputation quality for non-European populations compared to earlier European-centric panels. Notably, 58 (6%) of our mQTL index variants were insertions or deletions (indels), a class of variants that earlier imputation panels often failed to capture adequately. Indels can have substantial functional consequences but have been underrepresented in GWAS catalogs; their detection in our study reflects both improved imputation and the use of metabolomics as a sensitive intermediate phenotype.

Colocalization with the MVP phenome extends the catalog from genetic loci to candidate disease mechanisms. The implicated diseases consistently matched gene function: lipid-handling genes with dyslipidaemia, kidney-expressed transporters and enzymes with eGFR and chronic kidney disease, and urate-handling loci with gout. Pleiotropic hubs such as *FADS1*, *SLCO1B1* and *CPS1* connect dozens of metabolites to many trait families, consistent with disease effects that act through the pathways these genes control. Colocalization indicates a shared causal variant, however, not proof of a causal chain. *SLC16A9* illustrates this caveat. The locus links acylcarnitines to gout and coronary atherosclerosis, and a carnitine-mediated effect on cardiac lipotoxicity was plausible a priori. Mendelian randomization could not separate the carnitine-and urate-transport roles of *SLC16A9*, as expected for a single transporter carrying both substrates. Our estimate for urate on coronary artery disease resembles earlier Mendelian randomization work: robust with conventional estimators but attenuated under MR-Egger.^46^ Randomized trials of urate-lowering therapy have not reduced cardiovascular events in patients with ischemic heart disease^53^ or improved outcomes in heart failure.^54^ Nor did they slow atherosclerosis, measured either as carotid intima-media thickness^55^ or as coronary plaque volume.^56^ These trials started treatment late in life or after disease onset and ran for months to a few years, whereas genetic variants act lifelong. Additionally, these trials largely enrolled people without gout. In gout, reaching a serum urate target below 6 mg/dL was associated with fewer major adverse cardiovascular events^57^, but this setting cannot distinguish an effect of urate from suppression of gout-related inflammation, and the exposure was defined by treatment response rather than assignment. These results therefore do not exclude a contribution of urate to atherogenesis, but they argue against urate lowering as a cardiovascular intervention at least in the general population. We therefore do not interpret the coronary colocalization at *SLC16A9* as evidence of a modifiable urate-or carnitine-driven pathway. Genetically predicted carnitine has been associated with higher coronary artery disease risk^58^, but those instruments include pleiotropic transporter loci, so a carnitine-mediated pathway remains a hypothesis. We regard the locus as a robust statistical association whose precise causal pathway remains to be established.

*ENPEP* illustrates a different kind of insight. Fibrinopeptides have been proposed as a marker of disseminated intravascular coagulation^59,60^, but this has not become part of routine clinical practice, which instead typically measures fibrinogen or D-dimer directly. Fibrinopeptides have also been explored as a marker of other thrombotic conditions^61^, raising the possibility of broader future diagnostic use. Our results indicate that measured levels of these fragments are shaped not only by a patient’s coagulation state but also by their ACE-inhibitor treatment status and *ENPEP/ACE* genotype: ACE continues to act on the fibrinopeptide substrate pool generated during clot formation in the sample tube, so a patient’s ACE activity at the time of blood draw is captured alongside, and potentially confounded with, any coagulation-related signal. Should fibrinopeptide-based testing see wider clinical use, this would be a relevant consideration — and, conversely, these fragments could in principle serve as a biomarker of ACE-inhibitor activity itself. The findings more broadly connect the renin–angiotensin and coagulation systems.

Cross-study replication in metabolomics is bounded by platform comparability. NMR panels such as Nightingale, used in the largest mGWAS including UK Biobank, quantify about 250 predominantly lipoprotein, lipid, amino-acid and glycolysis-related measures, whereas our catalog rests on 1,955 untargeted Metabolon features that include xenobiotics, nucleotides, peptides and unnamed compounds, and UK Biobank has no urine metabolomics. Consistent with this, the NMR-based study supported only 4 of our 969 mQTLs and the untargeted MS study 73, together changing the classification of just 2 loci relative to the nine Metabolon studies when counting any biofluid and any ancestry as reported. Replication of urine and non-lipid signals therefore requires cohorts with the same platform and biofluid, which is why we combined GCKD with de-novo replication in CRIC.

Several limitations should be considered when interpreting our results. First, European American serum and urine sample sizes were unbalanced; the power-matched analysis shows that the European American biofluid-specificity proportions reflect power, and we therefore base quantitative biofluid comparisons on the balanced African American sample. Second, ARIC participants were older adults sampled at a single visit, ancestry groups were defined by self-report, and the cohort does not capture the full spectrum of human genetic diversity. Third, urine mQTLs among African Americans were replicated in CRIC, a chronic kidney disease cohort, whereas replication of African American serum signals relied on the European-ancestry GCKD study. Fourth, colocalization and Mendelian randomization support shared genetic effects rather than causality, and candidate causal genes and AlphaGenome predictions require experimental validation.

Our study demonstrates that multi-ancestry, multi-biofluid metabolomics GWAS designs substantially enhance genetic discovery and mechanistic interpretation. The paired analysis of serum and urine metabolomes revealed complementary genetic architecture, with approximately 75% of metabolites showing biofluid-specific associations in our balanced African American sample. Trans-ancestry finemapping improved resolution for both shared and population-specific signals, and colocalization with a clinical phenome connected prioritized enzymes and transporters to disease. The identification of an African American-enriched splice variant in *SLC28A1* illustrates how ancestral diversity can reveal functional variants with potential pharmacogenomic relevance.

As large-scale biobanks and metabolomics platforms continue to expand, integrating data across populations and biofluids will be essential for building a comprehensive atlas of human metabolic genetics. Our findings provide a template for such integrative approaches and highlight the scientific and equity imperatives for diversifying genomic research.

## Acknowledgements

The Atherosclerosis Risk in Communities study has been funded in whole or in part with Federal funds from the National Heart, Lung, and Blood Institute, National Institutes of Health, Department of Health, and Human Services, under Contract nos. (75N92022D00001, 75N92022D00002, 75N92022D00003, 75N92022D00004, 75N92022D00005). The authors thank the staff and participants of the ARIC study for their important contributions. Funding was also supported by R01HL087641 and R01HL086694; National Human Genome Research Institute contract U01HG004402; and National Institutes of Health contract HHSN268200625226C. Infrastructure was partly supported by Grant Number UL1RR025005, a component of the National Institutes of Health and NIH Roadmap for Medical Research. Visits 5 metabolomics measurements were sponsored in by grants R01HL141824 and 5R01DK124399. The CRIC metabolomics data was funded by P20GM109036-01A1 from the National Institute of General Medical Sciences. The work of P.S., I.S., and Z.R.H. was supported by the German Research Foundation (DFG) Project-ID 530592017 (SCHL 2292/3–1). The work of P.S., I.S., A.K. and N.S. was supported by the DFG Project-ID 499552394 – CRC 1597 Small Data. The work of P.S., A.K., A.W., and O.B. was supported by the DFG Project-ID 431984000 – CRC 1453 NephGen. The work of P.S. and A.K. was supported by the DFG Germany‘s Excellence Strategy (CIBSS – EXC-2189 – Project-ID 390939984). Dr. Coresh is supported by the Johns Hopkins O’Brien Center to Advance Kidney Health Equity (JHOC, 5U54DK137331). Drs. Li and Zhang are supported by R01DK135873 from the National Institute of Diabetes and Digestive and Kidney Disease.

## Author Contributions

Design of this study: P.S., B.Y.

Recruitment and management of study: E.B., Jo. C., M.G., T.K., Ji.C., J.H., C.L.

Genotyping: E.B.

Metabolite quantification: P.S., Jo.C., B.Y., M.G.

Bioinformatics and statistical analysis: I.S., N.S., A.S., Yo. L., O.B., S.M.M., R.Z., D.Z., Z.R.H, Yu. L., Q. N., P.S.

Interpretation of Results: I.S., A.W., A.K., M.G., B.Y., P.S.

Wrote the manuscript: I.S., P.S.

Critically read and approved the manuscript: All authors

## Methods

### Study design and participants

The Atherosclerosis Risk in Communities (ARIC) study is a prospective population-based cohort study with enrollment in 1987–1989, recruiting participants aged 45–64 years from four U.S. communities: Forsyth County, North Carolina; Jackson, Mississippi; suburbs of Minneapolis, Minnesota; and Washington County, Maryland.^24,25^ The study was designed to investigate the etiology and natural history of atherosclerosis and cardiovascular disease. Participants have undergone serial clinical examinations, with the collection of biological specimens at multiple time points.

The present study included ARIC participants with available genotype data and metabolomic profiling from visit 5 (2011–2013). Serum metabolomic profiling was available for all participants with available blood samples as well as consent while urine profiling was done in in a subsample that did not have previous kidney failure. This included all African Americans as well as European Americans sampled at the same sample size with oversampling of participants with eGFR<60 mL/min/1.73m^2^. Participants were stratified by self-reported race/ethnicity into European American (EA) and African American (AA) ancestry groups. After quality control, the final analytic sample comprised 4,431 participants with metabolite data: EA serum (N_serum_ = 3,607), EA urine (N_urine_ = 797), AA serum (N_serum_ = 818), and AA urine (N_urine_ = 794). The study was approved by the institutional review boards at each participating center, and all participants provided written informed consent (IRB00311861).

### Genotyping and imputation

Genotyping was performed using the Affymetrix Genome-Wide Human SNP Array 6.0 (Affymetrix, Santa Clara, CA). Standard quality control procedures were applied, including exclusion of variants if they had call rate <95%, minor allele frequency (MAF) <1%, or significant deviation from Hardy-Weinberg equilibrium (P < 1 × 10^−6^).

Genotype imputation was performed using the TOPMed Imputation Server (https://imputation.biodatacatalyst.nhlbi.nih.gov/) with the TOPMed r2 reference panel.^52^ Phasing was performed with Eagle v2.4 and imputation with Minimac4. Post-imputation, variants were filtered to retain those with imputation quality (R^2^) >0.3 and MAF >1% within each ancestry stratum. The TOPMed^21^ reference panel includes substantial representation of African individuals, enabling improved imputation accuracy and capture of small insertions and deletions (indels) that were poorly represented in earlier reference panels.

### Metabolite identification and quantification

Serum and urine metabolites were quantified using the Metabolon HD4 global metabolomics platform (Metabolon, Inc., Durham, NC).^62^ This untargeted approach employs ultrahigh-performance liquid chromatography–tandem mass spectrometry (UPLC-MS/MS) across four complementary ionization methods: two reverse-phase methods using positive ion mode electrospray ionization, one reverse-phase method using negative ion mode electrospray ionization, and one hydrophilic interaction liquid chromatography (HILIC) method using negative ion mode electrospray ionization.

Metabolites were identified by comparison of retention time, mass-to-charge ratio (m/z), and MS/MS fragmentation patterns to Metabolon’s proprietary library of authenticated chemical standards. Each metabolite was assigned a unique identifier and classified into super-pathways (e.g., amino acid, lipid, xenobiotics) and sub-pathways.

Raw metabolite abundance values were normalized to account for inter-day variation by rescaling each compound to set the median equal to 1 across run days. For serum metabolites, values were rank-based inverse normal transformed. For urinary metabolites, we applied probabilistic quotient (PQ) normalization to account for differences in urine concentration.^63^ Briefly, PQ factors were calculated as the median of the ratios of each metabolite’s abundance to a reference spectrum (median of all samples) across 372 endogenous metabolites with <1% missingness. Urinary metabolite values were divided by the sample-specific PQ factor, then rank-based inverse normal transformed. Metabolites with >50% missing values were excluded from analysis. Metabolites were retained for genome-wide association analysis if they had measurements in ≥300 participants within an ancestry-biofluid stratum.

The platform quantified a total of 1,316 metabolites in serum and 1,374 metabolites in urine after quality control. Metabolite annotations were harmonized across biofluids to reflect the newer (=urine) Metabolite annotation, yielding 1,955 unique analytes (581 serum-only, 639 urine-only, 735 detected in both biofluids).

### Definition of covariates

Covariates for GWAS were age, sex and the first 10 principal components of ancestry. Age was calculated as of the date of visit 5 examination. Sex was self-reported. Genetic principal components (PCs) 1-10 were derived from genome-wide genotype data using PLINK v1.9^64^ and calculated separately within each ancestry group.

### Genome-wide association analysis

Genome-wide association testing was performed separately within each ancestry-biofluid stratum (EA serum, EA urine, AA serum, AA urine) using REGENIE v3.1.^65^ REGENIE implements a two-step whole-genome regression approach that accounts for relatedness and population structure while maintaining computational efficiency for biobank-scale data. In step 1, a whole-genome regression model was fit using a subset of high-quality genotyped variants (MAF >1%, minor allele count > 100, genotyping rate >90%, Hardy-Weinberg equilibrium *P* > 1 × 10^−1^^5^). In step 2, association testing was performed for all imputed variants using a linear regression model with the step 1 predictions as an offset. All association tests were two-sided. The association model included the following covariates: age, sex, and the first 10 ancestry-specific principal components. Association testing was restricted to variants with MAF >1% and imputation R^2^ ≥0.3 within each stratum. Genomic inflation factors (λ) were calculated for each metabolite GWAS to assess residual population stratification or technical artifacts.

African American participants of ARIC show moderate, well-characterized European admixture (median proportion of European ancestry approximately 15%).^66^ Association testing within each ancestry stratum, adjustment for ten ancestry-specific genetic principal components and REGENIE’s leave-one-chromosome-out whole-genome regression, which captures relatedness and population structure,^65^ together account for the global-ancestry variation that admixture generates.

Study-wide significance was defined as *P* < 2.56 × 10^−1^^1^ (5 × 10^−8^ / 1,955 metabolites), corresponding to a Bonferroni correction for the number of unique metabolites tested. Metabolite quantitative trait loci (mQTLs) were defined by merging significant variants within ±500 kb windows within each ancestry and merging any overlapping windows (**Extended Data Fig. 2**).

Variance explained by index variants was compared across strata descriptively. Smaller discovery samples detect only larger-effect associations. Shared mQTLs were defined based on genomic proximity using a 1 Mb window. Specifically, a locus was considered shared across ancestries if the index variants for the same metabolite and biofluid in both the EA and AA analyses were located within 1 Mb of each other. Analogously, a locus was considered shared across biofluids if the index variants for the same metabolite in serum and urine were located within 1 Mb of each other. Per-stratum counts of mQTLs, distinct genomic regions, and metabolites with at least one genome-wide-significant association are provided in **Supplementary Table 5**. Loci that did not meet this proximity criterion were classified as biofluid-or candidate ancestry-specific, respectively. Each of the 632 candidate ancestry-specific mQTLs (590 EA-specific, 42 AA-specific; **Supplementary Tables 3-4**) was evaluated using a three-step rule to distinguish genuine ancestry-specific effects from those that are more parsimoniously explained as statistically driven, e.g. by imputation availability or power (**Extended Data Fig. 2**). First, availability: if the index variant, or a proxy at LD *r*² ≥ 0.8, was not available or did not pass quality control in the other ancestry, the locus was excluded as imputation-driven. Second, power: for loci passing the availability check, we calculated the power to detect the discovery effect size at the other ancestry’s observed sample size and standard error; loci with <80% power were excluded as power-driven. Third, heterogeneity: for the remaining loci, we tested for effect-size heterogeneity between ancestries using a two-sided z-test, applying a Bonferroni-corrected significance threshold (*P* < 0.05/632 = 7.9 × 10⁻⁵) to account for the number of loci tested; loci without significant heterogeneity were likewise excluded as statistically driven, and only those with significant heterogeneity were retained and classified as **ancestry-specific**. Full per-locus statistics (effect sizes and standard errors in both ancestries, power, and heterogeneity *P*-value) for the retained ancestry-specific loci are provided in **Supplementary Table 20**. At the level of statistical fine-mapping, a credible set is termed a fine-mapped ancestry-specific signal if MESuSiE assigns it to one ancestry (PIP > 0.5, with PIP_shared ≤ 0.5) in **Supplementary Tables 21 and 23**; this applies to every credible set in the dataset, independent of the locus-level classification above. Where an mQTL is independently classified as ancestry-specific at both levels, i.e., it passes the locus-level rule above and carries a fine-mapped ancestry-specific signal for the same ancestry, we refer to it as a fine-mapped ancestry-specific mQTL.

### Gene assignment

We prioritized candidate causal genes for each mQTL based on the ProGeM framework^67^ with additional extensions (https://github.com/genepi-freiburg/GWASannotation)^68^. The pipeline integrates gene proximity, LD-based gene overlap, functional variant consequences annotated with Ensembl VEP v101,^69^ genetic colocalization with *cis*-eQTL and *cis*-pQTL datasets, and polygenic priority scores (PoPS).^70^

For European American mQTLs, colocalization was performed against *cis*-eQTLs from GTEx v8^50^ and eQTLGen,^71^ and against *cis*-pQTLs from ARIC EA participants.^72^ For mQTLs among African Americans, *cis*-eQTL colocalization used the multi-ancestry Kachuri et al.^73^ eQTL resource, and *cis*-pQTL colocalization used *cis*-pQTLs from ARIC in AA participants.^72^ Colocalization was performed using the coloc.abf function from the coloc R package.^74^ We retained signals with evidence for a shared causal variant (PPH4 > 0.8) and suggestive *cis*-eQTL or *cis*-pQTL associations (−log₁₀(*P*) > 5).

Gene prioritization relied on a weighted scoring scheme. Proximity to the lead variant was scored as 1.00, 0.99, and 0.98 for the first, second, and third nearest genes, respectively. Genes overlapping the LD interval received 1.00, and VEP annotations indicating moderate or high impact contributed 1.02. *Cis*-eQTL association without evidence of colocalization contributed 0.5. Colocalization with *cis*-eQTLs in tissues of interest for metabolism (defined as tissues enriched in paired plasma and urine mGWAS findings^14^, GTEx v8^50^: adrenal gland, heart left ventricle, kidney cortex, liver, pancreas, small intestine terminal ileum, colon transverse, stomach; kidney-specific datasets: tubule and glomeruli) was scored at 1.04, while general *cis*-eQTL colocalization was scored at 1.03. For AA mQTLs, tissues of interest were not defined separately; colocalization with the Kachuri *cis*-eQTL resource was scored at 1.03 (general colocalization). Plasma *cis*-pQTL colocalization (ARIC PWAS^72^) was weighted at 1.05. PoPS scores were computed per locus and ancestry stratum; the gene with the highest PoPS score per locus received an additive weight of 0.5 and the second-highest 0.25.

### Covariate selection for multi-phenotype studies

To evaluate whether reported associations could reflect confounding by other correlated metabolites, we applied a method for covariate selection in multiphenotype studies (CMS)^75^ using the authors’ reference implementation (CMS_v1.0.R, function MC()), unmodified. For each of the 247 prioritized genes in each ancestry-by-biofluid stratum, we took the single most strongly associated (target metabolite, index variant) pair (366 gene-by-stratum combinations) and offered as candidate covariates the 50 metabolites most correlated with the target (|r| ≤ 0.8; measured in ≥90% of subjects with the target measured; remaining missingness mean-imputed). We derived an equivalent sample-size increase (SSincr) as the ratio of CMS-adjusted to marginal −log₁₀P (**Supplementary Table 12)**.

### Statistical finemapping

Multi-ancestry finemapping was performed using MESuSiE (Multi-ancestry Sum of Single Effects),^35^ which extends the Sum of Single Effects (SuSiE)^76^ model to jointly analyze GWAS summary statistics from multiple ancestries. MESuSiE explicitly models both shared and ancestry-specific causal signals using a mixture of bivariate normal distributions, allowing identification of variants with effects in one or both ancestries. For each mQTL region, we provided ancestry-specific GWAS summary statistics (effect estimates and standard errors) and LD matrices calculated from the ARIC genotype data.

For comparison, single-ancestry finemapping was performed using SuSiE^76^ separately within EA and AA strata based on the same set of shared variants. SuSiE decomposes the genetic signal into a sum of single-effect vectors, each representing one causal variant, and outputs posterior inclusion probabilities (PIPs) for each variant and 95% credible sets for each independent signal. Finemapping was performed for the genomic windows harboring mQTLs in serum and urine separately based on a harmonized cross-ancestry SNP set with joint variants. For shared loci, the window with the lowest *P* value across EA and AA was selected for joint multi-ancestry finemapping with MESuSiE. Multi-ancestry fine-mapping with MESuSiE was run on all 800 unique region-metabolite pairs (629 serum, 171 urine). For 6 of these (2 serum, 4 urine), MESuSiE returned no credible set, likely reflecting the absence of a remaining genome-wide significant variant (*P* < 5 × 10⁻⁸/1955 = 2.56 × 10⁻¹¹) within the harmonized cross-ancestry SNP set. Credible sets were therefore obtained for 794 region-metabolite pairs (627 serum, 167 urine). Single-ancestry finemapping with SuSiE was performed on the original per-ancestry windows, but based on the harmonized cross-ancestry SNP set to be comparable to MESuSiE. Multi-allelic variants were excluded from finemapping analyses to ensure consistent LD estimation.

Credible sets were classified as ancestry-shared (PIP_shared_ > 0.5), EA-specific (PIP_EA_ > 0.5 and PIP_shared_ ≤ 0.5), or AA-specific (PIP_AA_ > 0.5 and PIP_shared_ ≤ 0.5). Variants were considered high-confidence if their credible set contained ≤5 variants or their individual PIP exceeded 0.5. Finemapping resolution was assessed by credible set size (number of variants in the 95% credible set) and compared across methods and ancestries. Credible sets overlapping the MHC (major histocompatibility complex) region on chromosome 6 (25-34 Mb) are flagged in **Supplementary Tables 21 and 23** (column: ‘Located in MHC region’) and should be interpreted with caution, as the extended and complex LD structure in this region may pose challenges for statistical finemapping methods; MESuSiE’s original application to lipid traits excluded this region entirely.^35^

### Regional association plots

Regional association plots were generated in *R* (v4.3.2). For each finemapped region-metabolite pair, MeSuSiE PIPs and per-ancestry association statistics (−log10(*P*) derived from MeSuSiE *Z*-scores) were plotted across the locus interval. LD between each variant and the corresponding ancestry’s index variant (*r*²) was computed in the ARIC cohort and used for point coloring. Gene tracks were rendered with the *Gviz*^77^ *R* package using transcript annotation from TxDb.Hsapiens.UCSC.hg38.knownGene and gene symbols from org.Hs.eg.db, showing the longest transcript per gene.

### Variant annotation

Variant annotation was performed using the Ensembl Variant Effect Predictor (VEP) v101^69^ with the GRCh38 reference genome. Annotations included variant consequences (e.g., missense, splice-site, intronic), allele frequencies from gnomAD v3.1, and regulatory annotations from the ENCODE and Roadmap Epigenomics projects. Combined Annotation Dependent Depletion (CADD) scores^78^ were retrieved to prioritize potentially deleterious variants. VEP annotated 98.9% of finemapped variants with a predicted consequence.

Additionally, regulatory variant effects were predicted using AlphaGenome,^36^ a deep learning model that predicts the molecular consequences of genetic variants across multiple functional assays and cell types. For each finemapped variant, AlphaGenome scored the predicted impact on chromatin accessibility (ATAC-seq, DNase-seq), transcription factor binding (ChIP-seq), histone modifications, promoter activity (CAGE), and gene expression (RNA-seq), using a 1 Mb sequence context around each variant. Scores were computed across all available cell types and tissues from ENCODE or GTEx, with quantile scores reflecting the extremity of each predicted effect relative to common variants in gnomAD v3 (MAF > 1%). Variant-level summaries were generated by retaining the strongest predicted effect across all tracks, as well as separately for metabolically relevant tissues (defined as tissues enriched in paired plasma and urine mGWAS findings^14^: kidney, liver, adrenal gland, heart left ventricle, pancreas, small intestine terminal ileum, colon transverse, stomach). Variants with quantile scores exceeding 0.99 were considered to have reliably predicted functional effects and are reported as high-confidence, consistent with the model’s reported concordance with observed eQTL effect sizes at this threshold (Spearman r ≈ 0.73). In addition to the collapsed annotation in the supplementary tables, all individual annotations are provided in **Supplementary Data 2**. AlphaGenome annotation was performed using Python 3.12.3 and alphagenome v0.6.1. AlphaGenome predictions are in-silico, hypothesis-generating annotations of potential regulatory consequence; they were not experimentally validated in this study and are reported as supporting evidence for gene and variant prioritization.

### Population differentiation (Fst)

Genetic differentiation between European (CEU) and West African (YRI) reference populations was quantified using Fst, obtained from PopHuman’s^79^ precomputed Fst_CEU2YRI_10kb track, which reports values in non-overlapping 10-kb windows (hg19). Index variant positions were converted from GRCh38 to GRCh37/hg19. For 609 of the 969 (63%) mQTL index variants, the Fst value of the overlapping window was extracted using the rtracklayer R package, and expressed as a percentile relative to the genome-wide Fst distribution together with a one-sided empirical *P*-value (the proportion of genome-wide Fst values greater than or equal to the observed value). Index variants falling in a gap between windows have no Fst value, a property of the non-contiguous window track that was not further resolved; this applied to 37% of mQTLs and reflects window coverage rather than a systematic bias by locus type.

### Power-matched sensitivity analysis

To test whether apparent biofluid-specific biology in the European-American (EA) stratum reflects the larger serum sample size (N = 3,607) relative to urine (N = 797), we repeated the complete EA serum mGWAS restricted to the 797 EA participants who also contributed urine measurements. Gene set, pathway, and tissue enrichment analyses (GO, KEGG, GTEx tissue, and Metabolon super-/sub-pathway) were repeated on this power-matched locus set using the identical pipeline as the primary enrichment analysis.

### Pathway and tissue enrichment analyses

Gene set enrichment analysis was performed using the Kyoto Encyclopedia of Genes and Genomes (KEGG) pathway database.^80^ Enrichment was assessed using one-sided Fisher’s exact tests comparing the proportion of mQTL-implicated genes in each pathway to the genomic background. Pathways with fewer than 3 mapped genes were excluded. Fold enrichment was defined as the ratio of the fraction of mQTL-implicated genes in a pathway to the fraction of background genes in that pathway.

To identify tissue-specific gene sets for enrichment analyses, we used pre-compiled lists of the top 10% of specifically expressed genes across GTEx V8 tissues and human and murine kidney cell types, derived as described previously.^7^ Briefly, tissue-specific expression was assessed by ranking genes by t-statistics from a linear model comparing expression in each tissue or cell type to all others, following the approach of Finucane et al.^81^ The resulting top 10% gene lists per tissue were used as the basis for enrichment testing against our identified metabolite-associated genes. Gene set overrepresentation was tested analogously to KEGG. Of the 220 and 81 genes associated in serum and urine (overlap: 54), respectively, we were able to map 220 and 80 to the KEGG annotated genes. KEGG enrichment was performed using *clusterprofiler and KEGG.db v3.2.4*.

Pathway enrichment regarding annotated sub-and super-pathways as annotated by Metabolon Inc., was performed using Fisher’s exact tests.

In all enrichment analyses, statistical significance was determined by a Bonferroni-corrected significance threshold accounting for the number of pathways/gene sets in the background, respectively.

### Replication and cross-study comparisons

We assessed novelty of our 969 mQTLs against eleven published metabolite GWAS: nine Metabolon-annotated studies (Schlosser *et al.* (GCKD)^14^, Chen *et al.*^6^, Long *et al.*^4^, Yin *et al.*^20^, Hysi *et al.*^26^, Lotta *et al.*^27^, Surendran *et al.*^5^, Shin *et al.*^3^, Luo *et al.*^16^) and, as secondary-tier evidence, two studies on other platforms (Tambets *et al.*^2^, Nuclear Magnetic Resonance; Tahir *et al.*^18^, untargeted Mass Spectrometry) treated as a secondary tier. A locus was called known if the comparison study reported its own genome-wide-significant (P ≤ 5 × 10⁻⁸) index variant for the same metabolite within ±500 kb of the ARIC index variant, on the same chromosome, and all reported summary statistics are the comparison study’s own values for its own variant. Metabolite identity was matched using, for the primary tier, any concordant match among global Metabolon chemical ID, biofluid-specific chemical ID, or normalized biochemical name; for the secondary tier, normalized name only, supplemented for Tahir by an external-identifier bridge (PubChem, then KEGG, then HMDB; unambiguous 1:1 mappings only). Looking at biofluid-and ancestry-matched Metabolon-studies served as our primary novelty measure; counting a report in any biofluid and ancestry in all 11 studies as known served as a conservative bound. Full per-locus results across both definitions are provided in **Supplementary Table 9**. As a targeted replication analysis, we additionally evaluated the validation rate of our mQTLs against GCKD specifically — the only other study with a paired serum-and-urine design. For index variants not found directly in GCKD’s summary statistics, we searched for an LD proxy (*r*² ≥ 0.8, 1000 Genomes European reference panel: CEU, TSI, FIN, GBR, IBS) using LDlinkR, with allele phasing resolved via LDproxy’s correlated-alleles output, and used the proxy variant’s association statistics in its place. We then tested for concordant direction of effect at Bonferroni-corrected significance (*P* < 0.05/# lookups per biofluid) among all directly or proxy-matched pairs. Cross-ancestry and cross-biofluid consistency was assessed by comparing effect estimates between strata. Pearson correlation coefficients were calculated for effect sizes of shared mQTLs. For visualization, a regression slope was computed as the unweighted mean of the two per-direction regression slopes (regressing *y* on *x* and *x* on *y*), correcting for discovery-related winner’s curse.

As no published Metabolon urine resource exists for African American participants, we obtained de-novo replication in the Chronic Renal Insufficiency Cohort (CRIC), a prospective cohort of Black participants with mild-to-moderate chronic kidney disease recruited across seven U.S. clinical centers.^28,29^ Of 1,514 Black participants with baseline urinary metabolomics, 1,323 additionally had complete covariates and genome-wide genotyping and were included in this replication.

Urinary metabolites were profiled from baseline 24-hour urine samples on the same Metabolon UPLC-MS/MS platform as ARIC and quantified by area-under-the-curve against a library of authenticated standards. Metabolites were normalized using probabilistic quotient normalization: a reference set of metabolites with <50% missingness, excluding xenobiotics and unnamed compounds, was used to compute a per-sample scaling factor, followed by median-scaling across samples; each target metabolite was then rank-based inverse-normal transformed.

Genotyping was performed in 1,447 participants using the Illumina HumanOmni1-Quad BeadChip and imputed via the Michigan Imputation Server using the TOPMed r2 reference panel (GRCh38).

Covariates were age, sex, study site, and 10 genetic principal components; a sensitivity model additionally adjusted for eGFR (2021 CKD-EPI creatinine equation without race,^82^ log-transformed).

Of the 109 AA-urine mQTLs identified in ARIC (77 unique index variants, 101 unique metabolites), all 77 index variants were available in CRIC’s imputed genotype data at high quality (minimum Minimac R² = 0.950); 94 of 101 metabolites were measured in CRIC, yielding 102 testable variant-metabolite pairs. Association testing used REGENIE v3.2^65^ in two steps: a whole-genome regression model fitted on quality-controlled, LD-pruned genome-wide variants to generate leave-one-chromosome-out predictions for each metabolite, followed by extraction of the 77 index variants with PLINK 2 and single-variant association testing conditional on the step-1 predictions, under the primary and eGFR-adjusted models described above.

### Disease and complex-trait colocalization

We colocalized each of the four ancestry-by-biofluid mGWAS strata against the Million Veteran Program (MVP) phenome-wide GWAS^17^, testing European-ancestry ARIC strata against MVP European-ancestry summary statistics and African American ARIC strata against MVP African American summary statistics. We used the *genepicoloc R* package (https://github.com/genepi-freiburg/genepicoloc), which implements the colocalization approach from the coloc *R* package (coloc.abf^74^ with default priors). Regions in which colocalization was performed were defined as a ±500 kilobase window around the index variant (the variant with the lowest *P*-value in mGWAS, required to reach *P* < 5×10⁻⁸). We tested 1,669 traits (1,470 binary, 199 quantitative) from the European-ancestry MVP data and 1,368 traits (1,187 binary, 181 quantitative) from the African-ancestry MVP data; 3,037 trait-ancestry analyses in total. Colocalization was declared at a posterior probability of a shared causal variant (PP.H4) > 0.8; results were de-duplicated to one result per gene-metabolite-trait-family combination. Full per-hit results are provided in **Supplementary Table 25**.

### Medication-based observational analyses

To test whether pharmacologically reduced ACE activity alters fibrinopeptide fragment levels, we performed two individual-level observational analyses in ARIC. RASi medication use (ACE inhibitor, angiotensin-receptor blocker, and/or aldosterone antagonist in the past 4 weeks) was defined at Visit 5, versus no antihypertensive medication as the comparator; sensitivity analyses isolated each drug class individually and added a third, non-RASi-medication comparator group. Models were adjusted for age, sex, race, BMI, eGFR (CKD-EPI creatinine equation), diabetes medication use, smoking, and statin use, in the combined EA+AA sample (primary) and each ancestry separately. The first analysis tested seven clinically measured coagulation markers at Visit 1 (N = 4,122–4,209 per marker), with factor VII and fibrinogen also available at Visit 3 (N = 306); Visit 5 itself had no directly measured coagulation markers. These markers were therefore measured about two decades before medication ascertainment. The second analysis tested the same fibrinopeptide fragments plus aspartate, glycylvaline, aspartylaspartate, and serum potassium, all measured contemporaneously at Visit 5 (metabolite values rank-based inverse-normal transformed; potassium on its native scale). Associations were tested by multivariable linear regression.

### Mendelian randomization at SLC16A9

To determine whether SLC16A9’s colocalization with urate-related traits and coronary atherosclerosis (CAD) reflects one or two separable pathways, we performed two-sample MR for urate (CKDGen Round 4^83^, European ancestry) and five SLC16A9-linked acylcarnitines (carnitine, acetylcarnitine, propionylcarnitine, phenylacetylcarnitine, butyrylcarnitine; ARIC and Shin et al.^3^) against gout (CKDGen) and CAD (CARDIoGRAMplusC4D^84^). To test gout itself, rather than continuous urate, as a cause of CAD, we performed an additional MR using a case-control gout diagnosis GWAS (CKDGen R4) against CAD outcome with CARDIoGRAMplusC4D. For all analyses above, instrument SNPs were clumped using ieugwasr::ld_clump() against ancestry-matched 1000 Genomes Phase 3 reference panels^85^(r² < 0.001, 10 Mb window), and causal effects were estimated using inverse-variance-weighted regression, MR-Egger regression, and the weighted median estimator (or the Wald ratio for single-SNP instruments) via the TwoSampleMR R package.^86,87^

### Expression and splicing QTL lookup

For *SLC28A1*, we performed a lookup of the index variants *cis*-eQTL and *cis*-sQTL associations in GTEx v8^50^ across all 49 tissues. Significant associations were defined as *P* < 5 × 10^−8^.

### Generative AI language models

We employed generative AI language models, specifically Claude by Anthropic and ChatGPT by OpenAI, throughout the manuscript preparation process. These models facilitated iterative drafting and refinement of the content. However, all AI-generated contributions underwent thorough review, verification, and editing by the authors. Consequently, we hold complete responsibility for the accuracy, integrity, and originality of all content in the final manuscript.

## Data availability

Genome-wide summary statistics for all metabolite GWAS are deposited in the NHGRI-EBI GWAS Catalog (accession number: GCP001729; https://www.ebi.ac.uk/gwas/; release upon publication). Individual-level genotype and phenotype data from the ARIC study are available through dbGaP (accession phs000280.v7.p1) upon approval of data access requests. GTEx v8 data are available at https://gtexportal.org/.

## Code availability

We have clearly indicated each software whenever applicable and provided information on options (Methods). Code of the GWAS annotation pipeline is available under https://github.com/genepi-freiburg/GWASannotation. Statistical analyses were conducted using R v4.1.0 (Finemapping) or R v4.5.3 (remainder of the analyses). The following software packages were used: PLINK v1.9 and v2.0, REGENIE v3.1 (https://rgcgithub.github.io/regenie/), SuSiE/susieR v0.12.35 (https://stephenslab.github.io/susieR/), MESuSiE v1.0 (https://github.com/borangao/MESuSiE), VEP v101 (https://www.ensembl.org/vep), Python 3.12.3, AlphaGenome v0.6.1 ( https://github.com/google-deepmind/alphagenome), ProGeM (https://github.com/ds-cog-shg/ProGeM). Colocalization was performed using the *genepicoloc R* package (https://github.com/genepi-freiburg/genepicoloc). Visualization was performed using the *ggplot2*, *cowplot, patchwork* and *circlize* packages.

## References

1. Kettunen, J. et al. Genome-wide study for circulating metabolites identifies 62 loci and reveals novel systemic effects of LPA. Nat. Commun. 7, 11122 (2016).

2. Tambets, R. et al. Genetic analysis of circulating metabolic traits in 619,372 individuals. Nature 655, 971–978 (2026).

3. Shin, S. Y. et al. An atlas of genetic influences on human blood metabolites. Nat Genet 46, 543–550 (2014).

4. Long, T. et al. Whole-genome sequencing identifies common-to-rare variants associated with human blood metabolites. Nat Genet 49, 568–578 (2017).

5. Surendran, P. et al. Rare and common genetic determinants of metabolic individuality and their effects on human health. Nat Med 28, 2321–2332 (2022).

6. Chen, Y. et al. Genomic atlas of the plasma metabolome prioritizes metabolites implicated in human diseases. Nat Genet 10.1038/s41588-022-01270-1 (2023) doi:10.1038/s41588-022-01270-1.

7. Schlosser, P. et al. Genetic studies of urinary metabolites illuminate mechanisms of detoxification and excretion in humans. Nat. Genet. 52, 167–176 (2020).

8. Gieger, C. et al. Genetics meets metabolomics: a genome-wide association study of metabolite profiles in human serum. PLoS Genet. 4, e1000282 (2008).

9. Suhre, K. et al. A genome-wide association study of metabolic traits in human urine. Nat. Genet. 43, 565–569 (2011).

10. Kolz, M. et al. Meta-analysis of 28,141 individuals identifies common variants within five new loci that influence uric acid concentrations. PLoS Genet. 5, e1000504 (2009).

11. Yang, C. et al. Genomic atlas of the proteome from brain, CSF and plasma prioritizes proteins implicated in neurological disorders. Nat. Neurosci. 24, 1302–1312 (2021).

12. Reus, L. M. et al. Quantitative trait loci mapping of circulating metabolites in cerebrospinal fluid to uncover biological mechanisms involved in brain-related phenotypes. Mol. Psychiatry 30, 3478–3490 (2025).

13. Rhee, E. P. et al. A genome-wide association study of the human metabolome in a community-based cohort. Cell Metab. 18, 130–143 (2013).

14. Schlosser, P. et al. Genetic studies of paired metabolomes reveal enzymatic and transport processes at the interface of plasma and urine. Nat. Genet. 55, 995–1008 (2023).

15. Yang, C. et al. European and African ancestry-specific plasma protein-QTL and metabolite-QTL analyses identify ancestry-specific T2D effector proteins and metabolites. Nat. Commun. 16, 7412 (2025).

16. Luo, S. et al. Genome-wide association study of serum metabolites in the African American Study of Kidney Disease and Hypertension. Kidney Int. 100, 430–439 (2021).

17. Verma, A. et al. Diversity and scale: Genetic architecture of 2068 traits in the VA Million Veteran Program. Science 385, eadj1182 (2024).

18. Tahir, U. A. et al. Whole Genome Association Study of the Plasma Metabolome Identifies Metabolites Linked to Cardiometabolic Disease in Black Individuals. Nat. Commun. 13, 4923 (2022).

19. Yousri, N. A. et al. Whole-exome sequencing identifies common and rare variant metabolic QTLs in a Middle Eastern population. Nat. Commun. 9, 333 (2018).

20. Yin, X. et al. Genome-wide association studies of metabolites in Finnish men identify disease-relevant loci. Nat. Commun. 13, 1644 (2022).

21. Taliun, D. et al. Sequencing of 53,831 diverse genomes from the NHLBI TOPMed Program. Nature 590, 290–299 (2021).

22. Sengupta, D. et al. Performance and accuracy evaluation of reference panels for genotype imputation in sub-Saharan African populations. Cell Genomics 3, 100332 (2023).

23. McCarthy, S. et al. A reference panel of 64,976 haplotypes for genotype imputation. Nat. Genet. 48, 1279–1283 (2016).

24. 24. The Atherosclerosis Risk in Communities (ARIC) Study: design and objectives. The ARIC investigators. Am. J. Epidemiol. 129, 687–702 (1989).

25. Wright, J. D. et al. The ARIC (Atherosclerosis Risk In Communities) Study: JACC Focus Seminar 3/8. J. Am. Coll. Cardiol. 77, 2939–2959 (2021).

26. Hysi, P. G. et al. Metabolome Genome-Wide Association Study Identifies 74 Novel Genomic Regions Influencing Plasma Metabolites Levels. Metabolites 12, (2022).

27. Lotta, L. A. et al. A cross-platform approach identifies genetic regulators of human metabolism and health. Nat Genet 53, 54–64 (2021).

28. Feldman, H. I. et al. The Chronic Renal Insufficiency Cohort (CRIC) Study: Design and Methods. J Am Soc Nephrol 14, S148–53 (2003).

29. Lash, J. P. et al. Chronic Renal Insufficiency Cohort (CRIC) Study: baseline characteristics and associations with kidney function. Clin J Am Soc Nephrol 4, 1302–11 (2009).

30. Manczinger, M. et al. Pathogen diversity drives the evolution of generalist MHC-II alleles in human populations. PLOS Biol. 17, e3000131 (2019).

31. Dyer, S. C. et al. Ensembl 2025. Nucleic Acids Res. 53, D948–D957 (2025).

32. Persaud, A. K. et al. Increased renal elimination of endogenous and synthetic pyrimidine nucleosides in concentrative nucleoside transporter 1 deficient mice. Nat. Commun. 14, 3175 (2023).

33. Wevers, R. A. et al. Functional disruption of pyrimidine nucleoside transporter CNT1 results in a novel inborn error of metabolism with high excretion of uridine and cytidine. J. Inherit. Metab. Dis. 42, 494–500 (2019).

34. Khan, A. et al. Machine-learning-guided discovery of SLC25A45 as a mediator of mitochondrial methylated amino acid import and carnitine synthesis. Cell Metab. 37, 2220–2232.e8 (2025).

35. Gao, B. & Zhou, X. MESuSiE enables scalable and powerful multi-ancestry fine-mapping of causal variants in genome-wide association studies. Nat. Genet. 56, 170–179 (2024).

36. Avsec, Ž., et al. Advancing regulatory variant effect prediction with AlphaGenome. Nature 649, 1206–1218 (2026).

37. Lau, H. H., Ng, N. H. J., Loo, L. S. W., Jasmen, J. B. & Teo, A. K. K. The molecular functions of hepatocyte nuclear factors -In and beyond the liver. J. Hepatol. 68, 1033–1048 (2018).

38. Wallerman, O. et al. Molecular interactions between HNF4a, FOXA2 and GABP identified at regulatory DNA elements through ChIP-sequencing. Nucleic Acids Res. 37, 7498–7508 (2009).

39. Ramji, D. P. & Foka, P. CCAAT/enhancer-binding proteins: structure, function and regulation. Biochem. J. 365, 561–575 (2002).

40. Kaczynski, J., Cook, T. & Urrutia, R. Sp1-and Krüppel-like transcription factors. Genome Biol. 4, 206 (2003).

41. Gheorghe, M. et al. A map of direct TF–DNA interactions in the human genome. Nucleic Acids Res. 47, e21–e21 (2019).

42. Puig, R. R., Boddie, P., Khan, A., Castro-Mondragon, J. A. & Mathelier, A. UniBind: maps of high-confidence direct TF-DNA interactions across nine species. BMC Genomics 22, 482 (2021).

43. Xu, X. et al. Genetic imputation of kidney transcriptome, proteome and multi-omics illuminates new blood pressure and hypertension targets. Nat. Commun. 15, 2359 (2024).

44. 44. Jallow, M. W. et al. Metabolomics Ex/GWAS in the Amish reveals novel insights into cardiometabolic disease pathways. Preprint at 10.1101/2025.08.04.667775 (2025).

45. Suhre, K. et al. Human metabolic individuality in biomedical and pharmaceutical research. Nature 477, 54–60 (2011).

46. White, J. et al. Plasma urate concentration and risk of coronary heart disease: a Mendelian randomisation analysis. Lancet Diabetes Endocrinol. 4, 327–336 (2016).

47. Caplin, B. et al. Alanine-glyoxylate aminotransferase-2 metabolizes endogenous methylarginines, regulates NO, and controls blood pressure. Arterioscler. Thromb. Vasc. Biol. 32, 2892–2900 (2012).

48. Hu, X.-L. et al. Considerable impacts of AGXT2 V140I polymorphism on chronic heart failure in the Chinese population. Atherosclerosis 251, 255–262 (2016).

49. Gray, J. H., Owen, R. P. & Giacomini, K. M. The concentrative nucleoside transporter family, SLC28. Pflugers Arch. 447, 728–734 (2004).

50. GTEx Consortium et al. Genetic effects on gene expression across human tissues. Nature 550, 204–213 (2017).

51. Pastor-Anglada, M., Cano-Soldado, P., Errasti-Murugarren, E. & Casado, F. J. SLC28 genes and concentrative nucleoside transporter (CNT) proteins. Xenobiotica Fate Foreign Compd. Biol. Syst. 38, 972–994 (2008).

52. Das, S. et al. Next-generation genotype imputation service and methods. Nat. Genet. 48, 1284–1287 (2016).

53. Mackenzie, I. S. et al. Allopurinol versus usual care in UK patients with ischaemic heart disease (ALL-HEART): a multicentre, prospective, randomised, open-label, blinded-endpoint trial. The Lancet 400, 1195–1205 (2022).

54. Givertz, M. M. et al. Effects of Xanthine Oxidase Inhibition in Hyperuricemic Heart Failure Patients: The Xanthine Oxidase Inhibition for Hyperuricemic Heart Failure Patients (EXACT-HF) Study. Circulation 131, 1763–1771 (2015).

55. Tanaka, A. et al. Febuxostat does not delay progression of carotid atherosclerosis in patients with asymptomatic hyperuricemia: A randomized, controlled trial. PLOS Med. 17, e1003095 (2020).

56. Yu, M. et al. No evidence of coronary plaque stabilization by allopurinol in patients with acute coronary syndrome. J. Cardiovasc. Comput. Tomogr. 18, 195–202 (2024).

57. Cipolletta, E. et al. Treat-to-Target Urate-Lowering Treatment and Cardiovascular Outcomes in Patients With Gout. JAMA Intern. Med. 186, 332 (2026).

58. Zhao, J. V., Burgess, S., Fan, B. & Schooling, C. M. l-carnitine, a friend or foe for cardiovascular disease? A Mendelian randomization study. BMC Med. 20, 272 (2022).

59. Bick, R. L. & Baker, W. F. Diagnostic efficacy of the D-Dimer assay in disseminated intravascular coagulation (DIC). Thromb. Res. 65, 785–790 (1992).

60. Boisclair, M. D., Ireland, H. & Lane, D. A. Assessment of hypercoagulable states by measurement of activation fragments and peptides. Blood Rev. 4, 25–40 (1990).

61. Eisenberg, P. R. et al. Fibrinopeptide A levels indicative of pulmonary vascular thrombosis in patients with primary pulmonary hypertension. Circulation 82, 841–847 (1990).

62. Ford, L. et al. Precision of a Clinical Metabolomics Profiling Platform for Use in the Identification of Inborn Errors of Metabolism. J. Appl. Lab. Med. 5, 342–356 (2020).

63. Dieterle, F., Ross, A., Schlotterbeck, G. & Senn, H. Probabilistic quotient normalization as robust method to account for dilution of complex biological mixtures. Application in 1H NMR metabonomics. Anal. Chem. 78, 4281–4290 (2006).

64. Chang, C. C. et al. Second-generation PLINK: rising to the challenge of larger and richer datasets. GigaScience 4, 7 (2015).

65. Mbatchou, J. et al. Computationally efficient whole-genome regression for quantitative and binary traits. Nat. Genet. 53, 1097–1103 (2021).

66. Cheng, C.-Y. et al. Admixture mapping of obesity-related traits in African Americans: the Atherosclerosis Risk in Communities (ARIC) Study. Obesity 18, 563–572 (2010).

67. Stacey, D. et al. ProGeM: a framework for the prioritization of candidate causal genes at molecular quantitative trait loci. Nucleic Acids Res. 47, e3 (2019).

68. Monteiro-Martins, S. et al. Genetic screens of imaging-derived kidney volumes identify genes linked to kidney function. Kidney Int. 109, 365–378 (2026).

69. McLaren, W. et al. The Ensembl Variant Effect Predictor. Genome Biol. 17, 122 (2016).

70. Weeks, E. M. et al. Leveraging polygenic enrichments of gene features to predict genes underlying complex traits and diseases. Nat. Genet. 55, 1267–1276 (2023).

71. Võsa, U. et al. Large-scale cis-and trans-eQTL analyses identify thousands of genetic loci and polygenic scores that regulate blood gene expression. Nat. Genet. 53, 1300–1310 (2021).

72. Zhang, J. et al. Plasma proteome analyses in individuals of European and African ancestry identify cis-pQTLs and models for proteome-wide association studies. Nat. Genet. 54, 593–602 (2022).

73. Kachuri, L. et al. Gene expression in African Americans, Puerto Ricans and Mexican Americans reveals ancestry-specific patterns of genetic architecture. Nat. Genet. 55, 952– 963 (2023).

74. Giambartolomei, C. et al. Bayesian test for colocalisation between pairs of genetic association studies using summary statistics. PLoS Genet. 10, e1004383 (2014).

75. Aschard, H. et al. Covariate selection for association screening in multiphenotype genetic studies. Nat. Genet. 49, 1789–1795 (2017).

76. Wang, G., Sarkar, A., Carbonetto, P. & Stephens, M. A simple new approach to variable selection in regression, with application to genetic fine mapping. J. R. Stat. Soc. Ser. B Stat. Methodol. 82, 1273–1300 (2020).

77. Hahne, F. & Ivanek, R. Visualizing Genomic Data Using Gviz and Bioconductor. Methods Mol. Biol. 1418, 335–351 (2016).

78. Rentzsch, P., Witten, D., Cooper, G. M., Shendure, J. & Kircher, M. CADD: predicting the deleteriousness of variants throughout the human genome. Nucleic Acids Res. 47, D886– D894 (2019).

79. Casillas, S. et al. PopHuman: the human population genomics browser. Nucleic Acids Res. 46, D1003–D1010 (2018).

80. Kanehisa, M., Furumichi, M., Sato, Y., Matsuura, Y. & Ishiguro-Watanabe, M. KEGG: biological systems database as a model of the real world. Nucleic Acids Res. 53, D672–D677 (2025).

81. Finucane, H. K. et al. Heritability enrichment of specifically expressed genes identifies disease-relevant tissues and cell types. Nat. Genet. 50, 621–629 (2018).

82. Inker, L. A. et al. New Creatinine-and Cystatin C-Based Equations to Estimate GFR without Race. N Engl J Med 385, 1737–1749 (2021).

83. Tin, A. et al. Target genes, variants, tissues and transcriptional pathways influencing human serum urate levels. Nat. Genet. 51, 1459–1474 (2019).

84. the CARDIoGRAMplusC4D Consortium. A comprehensive 1000 Genomes–based genome-wide association meta-analysis of coronary artery disease. Nat. Genet. 47, 1121–1130 (2015).

85. Siva, N. 1000 Genomes project. Nat. Biotechnol. 26, 256 (2008).

86. Hemani, G., Tilling, K. & Davey Smith, G. Orienting the causal relationship between imprecisely measured traits using GWAS summary data. PLOS Genet. 13, e1007081 (2017).

87. Hemani, G. et al. The MR-Base platform supports systematic causal inference across the human phenome. eLife 7, e34408 (2018).

