## Supplementary Material for "Genetic effects on serum and urine metabolites across ancestries and their links to disease"

**Extended Data Figure 1: Overview of the multi-ancestry metabolomics genome-wide association study (mGWAS) workflow.**

Paired serum and urine mGWAS were conducted in European American (EA, $N_{serum}$ = 3,607, $N_{urine}$ = 797) and African American (AA, $N_{serum}$ = 818, $N_{urine}$ = 794) participants of the Atherosclerosis Risk in Communities (ARIC) study using 16,467,669 variants imputed via the Trans-Omics for Precision Medicine (TOPMed) reference panel and 1,955 unique metabolites measured on the Metabolon HD4 platform. Downstream analyses included integrated multi-ancestry comparisons with multi-ancestry finemapping via MESuSIE and cross-cohort replication in the German Chronic Kidney Disease (GCKD) and Chronic Renal Insufficiency Cohort (CRIC) study, multi-biofluid comparison of serum and urine mQTLs, and mQTL characterization through gene annotation, variant effect prediction (VEP), AlphaGenome variant annotation, and enrichment analyses using Kyoto Encyclopedia of Genes and Genomes (KEGG), and Genotype-Tissue Expression (GTEx) V8 databases as well as metabolite pathways as annotated by Metabolon. MAF: minor allele frequency. Created in BioRender. Schlosser, P. (2026) https://BioRender.com/o32v15a.

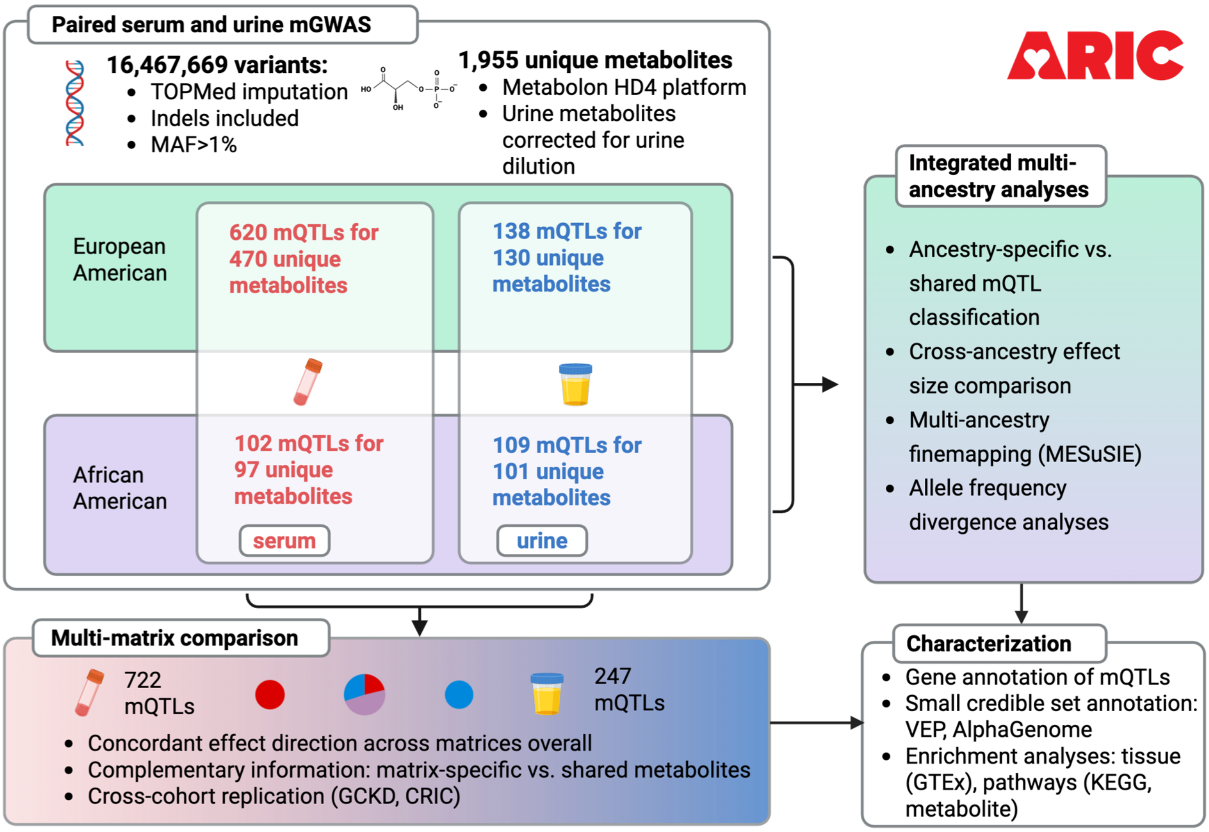

**Extended Data Figure 2: Terminology and classification rules for mQTL, biofluid-specificity, and ancestry-specificity.**

**(a) mQTL definition.** Genome-wide-significant variants are merged into one mQTL within ±500 kb, with overlapping windows merged. The merged mQTL's index SNP is its lowest-P variant; a second, independent signal may only be resolved later via fine-mapping. **(b) Biofluid-specificity.** A locus is biofluid-shared if the serum and urine index variants lie within 1 Mb of each other; otherwise it is biofluid-specific. Faded circles indicate no genome-wide-significant variant in that biofluid. **(c) Ancestry-specificity: two-tier definition.** *Tier 1:* analogous to (b), applied across ancestries, shared if index variants lie within 1 Mb, otherwise a candidate ancestry-specific mQTL. *Tier 2:* MESuSiE fine-mapping independently classifies credible sets as ancestry-shared or as a fine-mapped ancestry-specific signal, based on posterior inclusion probabilities. *Right:* a sequential rule excludes candidate ancestry-specific mQTLs that are better explained by incomplete testability or insufficient power.

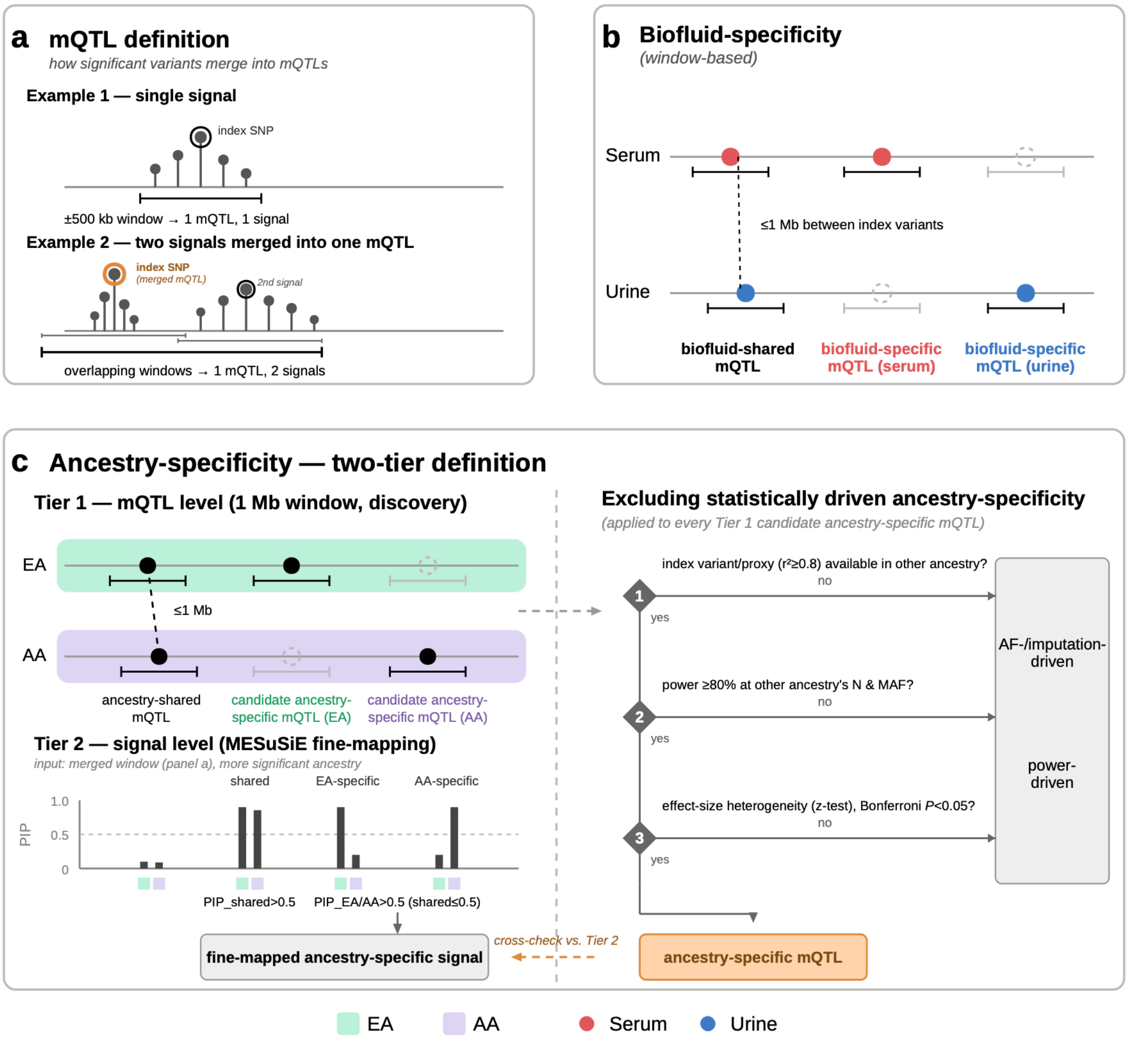

**Extended Data Figure 3: Overview of European American metabolite quantitative trait loci (mQTL), across biofluids.**

The left pie chart represents metabolites with European American (EA) mQTLs that were exclusively measured in serum, the right pie chart represents metabolites exclusively measured in urine, and the central pie chart represents metabolites measured in both biofluids, subdivided into those with serum mQTLs only (red), urine mQTLs only (blue), and mQTLs in both biofluids (purple). Pie chart sizes are proportional to the number of metabolites in each category. Metabolites without any EA mQTL are omitted. In total, 735 metabolites were measured in both biofluids, 639 in urine only and 581 in serum only. EA sample sizes were 3,607 in serum and 797 in urine.

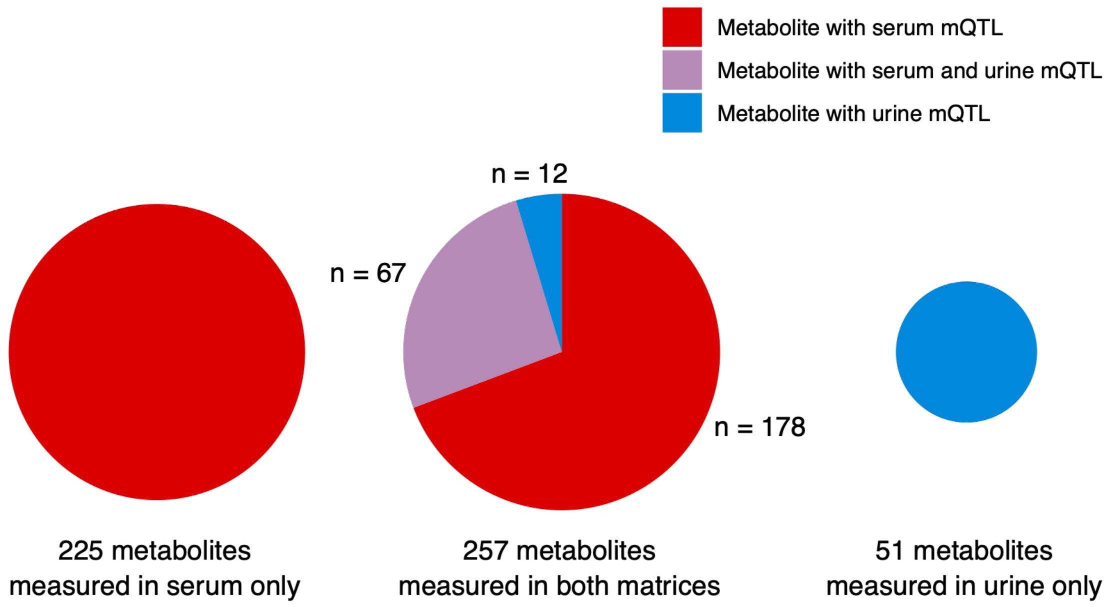

**Extended Data Figure 4: Overview of European American metabolite quantitative trait loci (mQTL) in the power-matched sensitivity analyses, across biofluids.**

The left pie chart represents metabolites with European American (EA) mQTLs that were exclusively measured in serum, the right pie chart represents metabolites exclusively measured in urine, and the central pie chart represents metabolites measured in both biofluids, subdivided into those with serum mQTLs only (red), urine mQTLs only (blue), and mQTLs in both biofluids (purple). Pie chart sizes are proportional to the number of metabolites in each category. Metabolites without any EA mQTL are omitted. In total, 735 metabolites were measured in both biofluids, 639 in urine only and 581 in serum only. EA sample size in the power-matched analyses was 797 in serum and urine.

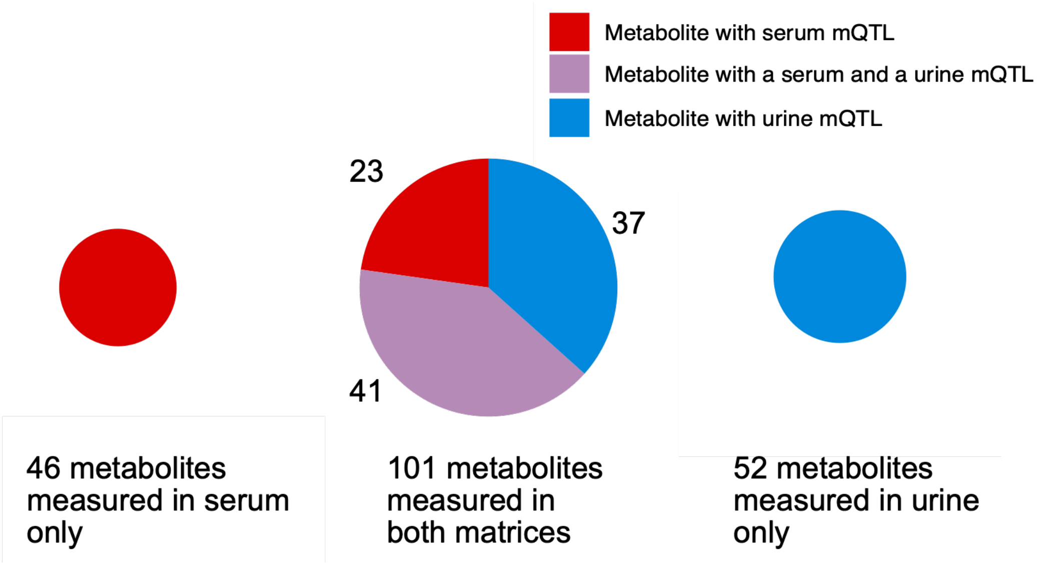

**Extended Data Figure 5: Replication of ARIC metabolite quantitative trait locus (mQTL) effect sizes in the German Chronic Kidney Disease (GCKD) study.**

Each point represents an mQTL, with error bars indicating ±1.96*standard error. Points are colored by the $-\log_{10}(P)$ of the ARIC discovery association. The dashed gray line represents the identity line, and the solid red line shows the linear regression fit. Pearson correlation coefficients, regression slopes, and the number of matched mQTLs are annotated in each panel. (a) EA ARIC serum mQTLs in GCKD plasma. (b) AA ARIC serum mQTLs in GCKD plasma. (c) EA ARIC urine mQTLs in GCKD urine. (d) AA ARIC urine mQTLs in GCKD urine.

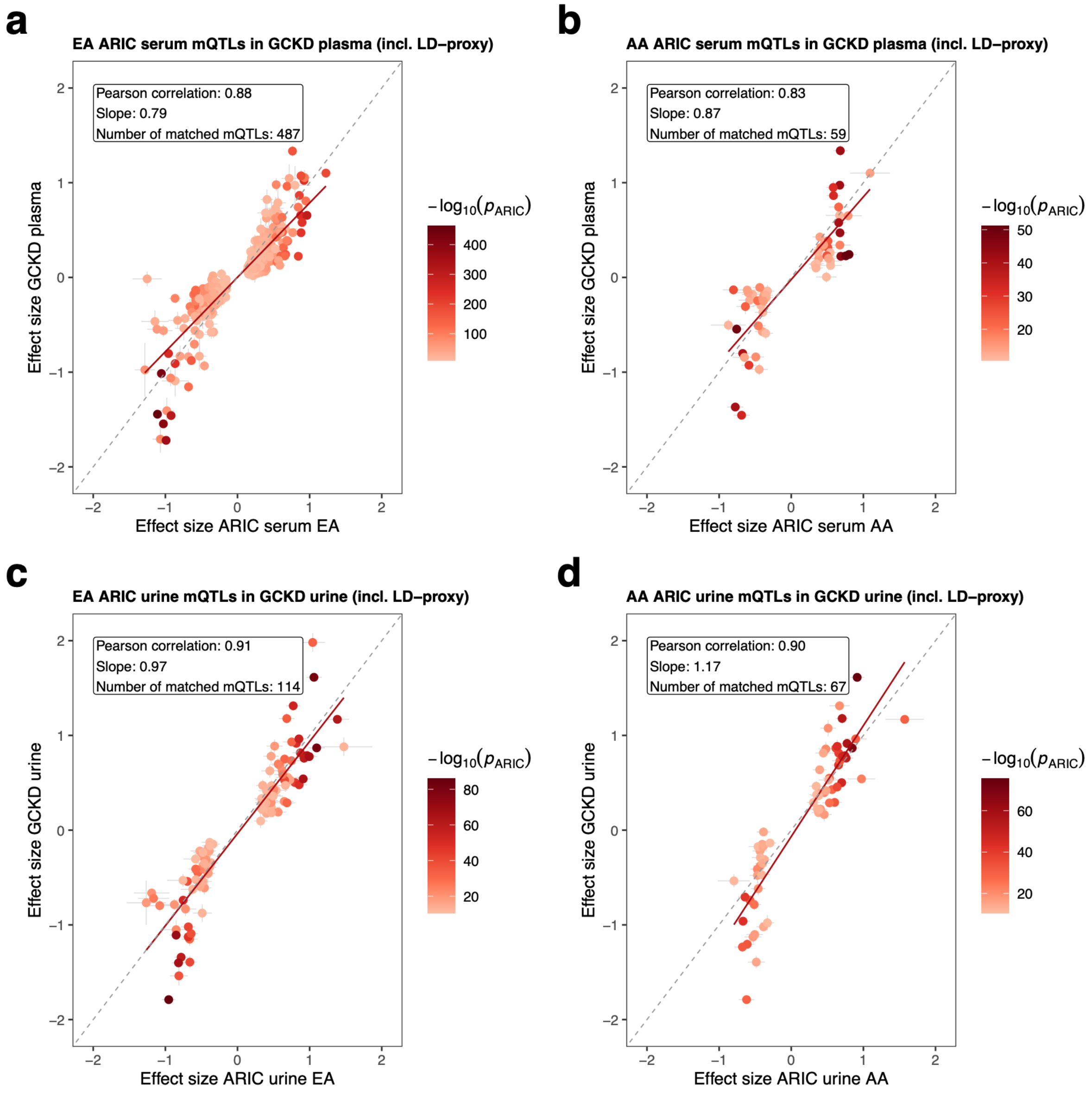

**Extended Data Figure 6: Functional classification of the 247 prioritized candidate causal genes for metabolite quantitative trait loci (mQTLs) in ARIC.**

Pie chart showing the distribution of enzyme classes and transporter annotations across the 247 unique genes prioritized by the ProGeM-based framework integrating proximity, linkage disequilibrium overlap, VEP consequences, cis-eQTL and cis-pQTL colocalization, and Polygenic Priority Score (PoPS).

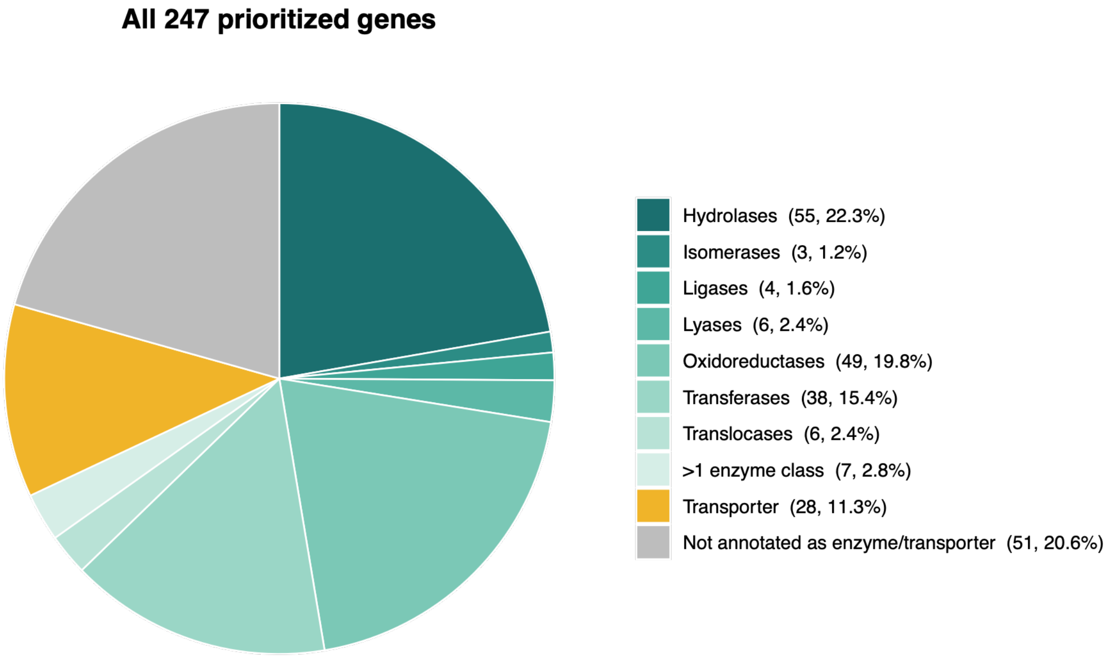

**Extended Data Figure 7: Allele frequency comparison of mQTL index variants between EA and AA in serum.**

Minor allele frequencies in EA (x-axis) versus AA (y-axis) for shared index variants in serum, with marginal histograms displaying the frequency distributions for candidate ancestry-specific variants (red: AA-specific; blue: EA-specific). Circles mark variants where the same allele is the minor allele in both ancestries; triangles mark variants where this differs. Trend lines (LOESS) are shown separately for EA-associated and AA-associated mQTLs. Gray crosses mark variants absent from the opposing ancestry.

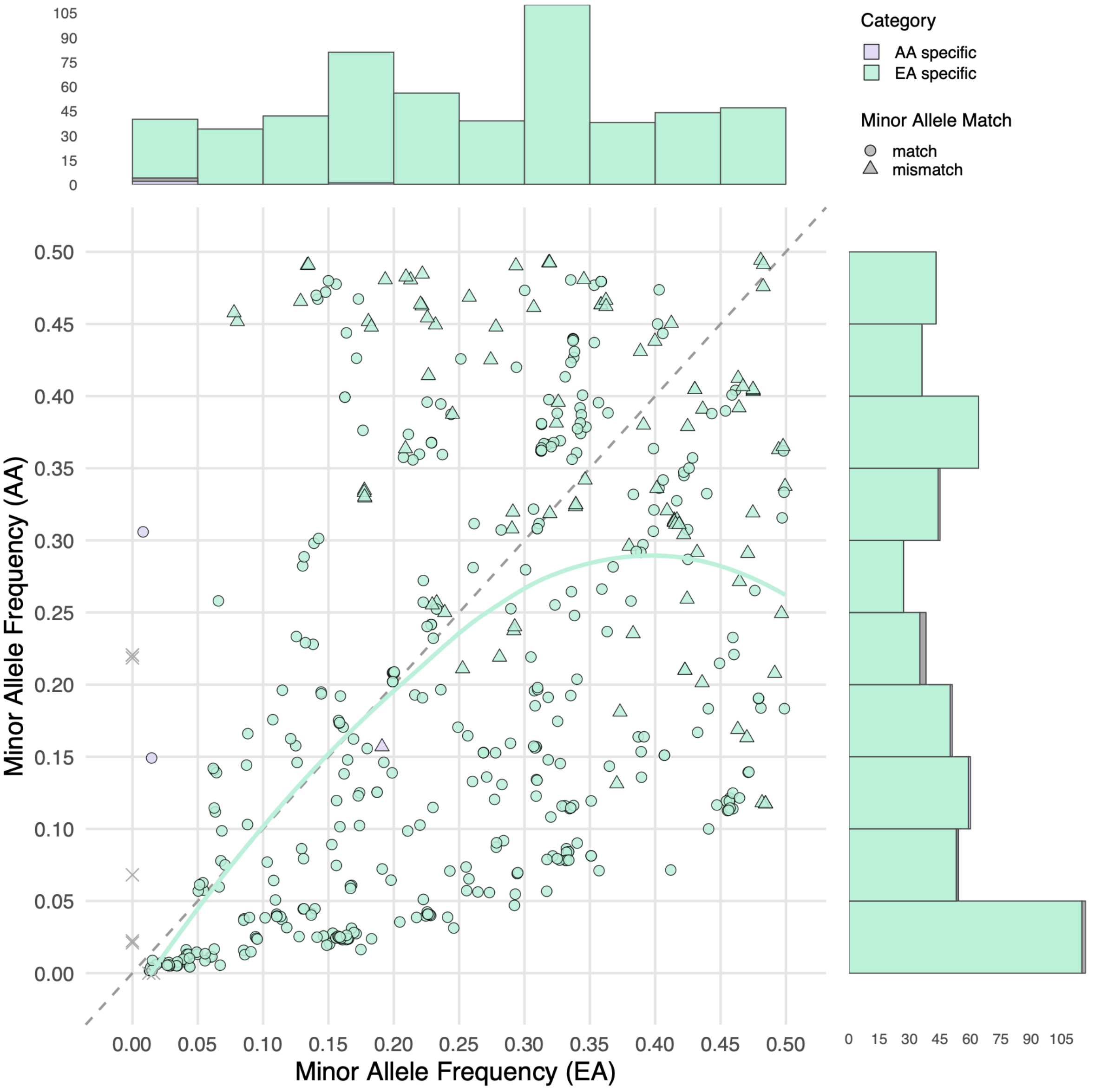

**Extended Data Figure 8: Heatmap comparing the number of independent signals in serum identified per region by MESuSiE (multi-ancestry fine-mapping) versus single-ancestry SuSiE analyses in African American (AA) (a) and European American (EA) (b).**

Circle size reflects the number of regions at each combination; circles above the diagonal (MESuSiE ≥ SuSiE) indicate regions where multi-ancestry fine-mapping detected at least as many signals (credible sets) as single-ancestry analysis.

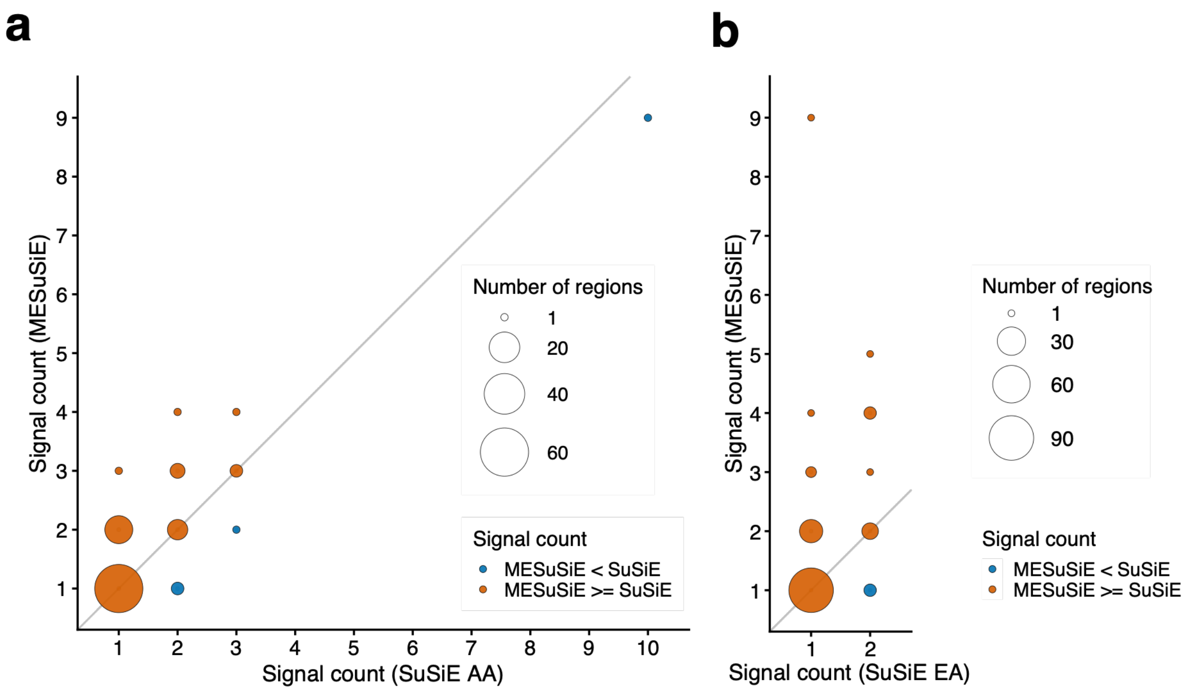

**Extended Data Figure 9: AlphaGenome-predicted regulatory effects of variant chr11:61820833:A>G at the FADS1–FADS2 locus.**

Shown are AlphaGenome difference scores (alternative minus reference allele) across the 1 Mb prediction window (including tolerance) centered on the variant (orange vertical line). The top panel displays gene annotations. Subsequent panels show predicted effects on transcription factor ChIP-seq occupancy (HNF4A, RXRA) in liver, DNase-seq chromatin accessibility in liver and kidney, and CAGE transcription start site activity on both strands in liver and kidney. Gray shading at the variant position indicates the predicted allelic effect at the variant itself, while colored tracks show the predicted effects across the surrounding sequence context. The variant is located between FADS1 and FADS2 and shows predicted effects on HNF4A and RXRA binding as well as chromatin accessibility in kidney at the variant position.

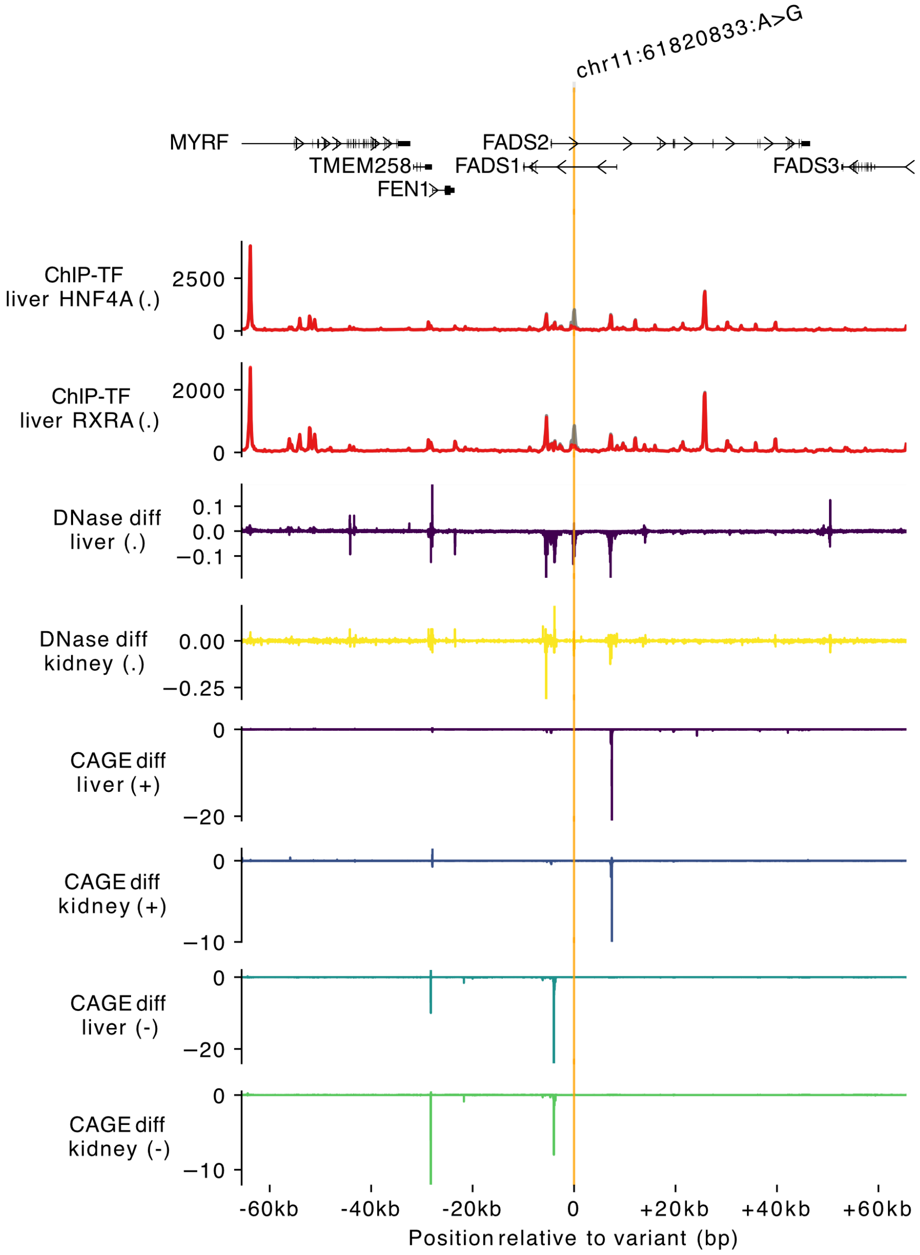

**Extended Data Figure 10: Details on the *ALDH7A1* locus.**

**a) Regional association plot of the ALDH7A1 locus.** From top to bottom: (1) Posterior Inclusion Probabilities (PIPs) from MeSuSiE multi-ancestry fine-mapping. Variants belonging to credible sets are colored by credible set membership (L1, L2, etc.); remaining variants are shown in gray. The EA index variant is indicated by a purple circle and the AA index variant by an orange triangle. (2, 3) Regional association in the European- and African-American-ancestry populations, respectively, showing −log10(*P*) derived from MeSuSiE Z-scores. Points are colored by $r^2$ with the corresponding ancestry's index variant (gray: no LD information).
**b) AlphaGenome-predicted variant effects at the ALDH7A1 locus (chr5:126595258:C>T, rs144272515).** Top: gene annotation showing ALDH7A1 and neighboring PHAX. The variant position is indicated by the vertical orange line. Panels show the predicted effect of the alternate allele: increased activity of a promoter-associated histone mark in three tissues (blood, liver, kidney), and predicted chromatin accessibility. Horizontal axis shows position in base pairs relative to the variant.

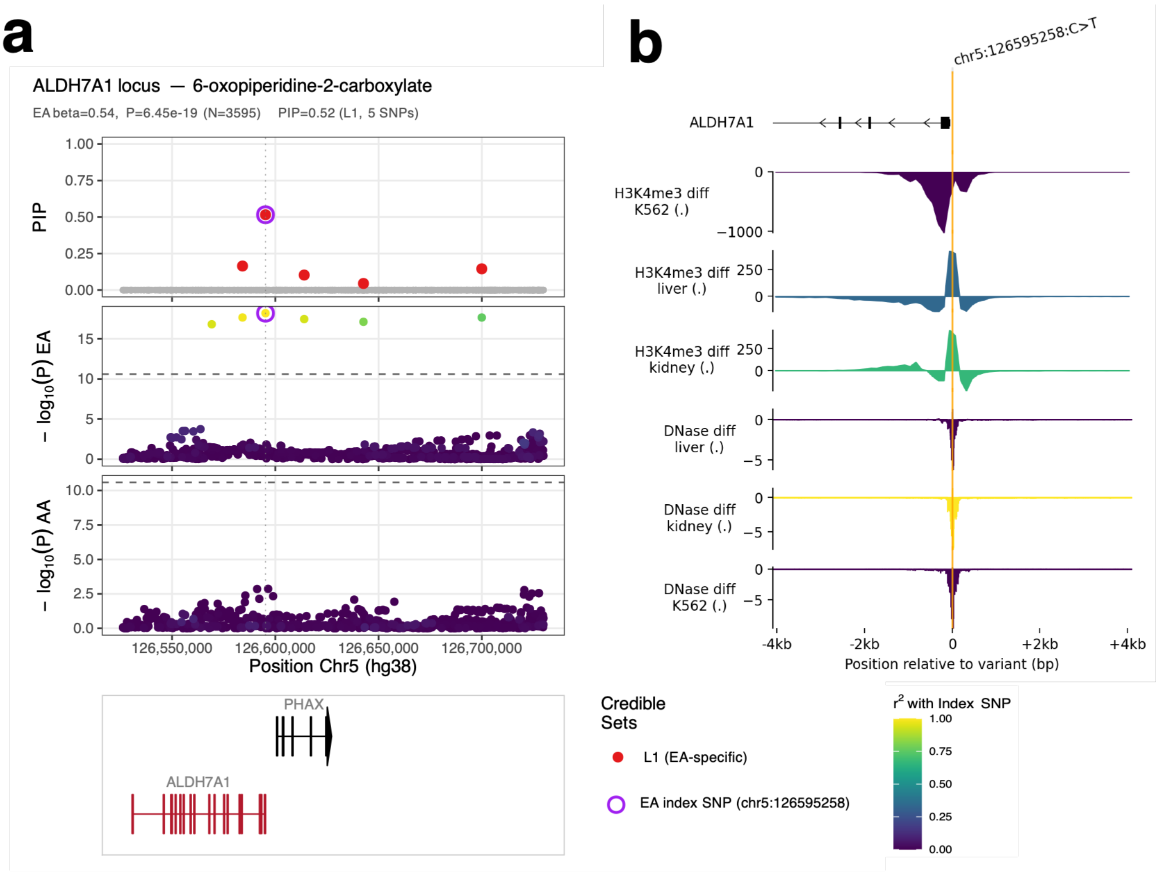

**Extended Data Figure 11: Details on the *ENPEP/ACE* locus showcase.**

**Schematic of the proposed *ENPEP/ACE* link between blood pressure regulation and fibrinopeptide degradation.** The upper panel shows thrombin-mediated release of fibrinopeptides A and B from fibrinogen and their subsequent degradation. Numbers indicate amino acid positions. The lower panel shows the renin–angiotensin system. *ENPEP* and *ACE* act in both pathways. The green icon and dashed line mark colocalization of *ENPEP*-associated fibrinopeptide A fragments with blood pressure traits and potassium. The solid box summarizes follow-up analyses in ARIC individual participant data. ACE, angiotensin-converting enzyme; ANPEP, aminopeptidase N; APTT, activated partial thromboplastin time; ARIC, Atherosclerosis Risk in Communities study; BP, blood pressure; CPB2, carboxypeptidase B2; ENPEP, glutamyl aminopeptidase (aminopeptidase A); ERAP1, endoplasmic reticulum aminopeptidase 1.

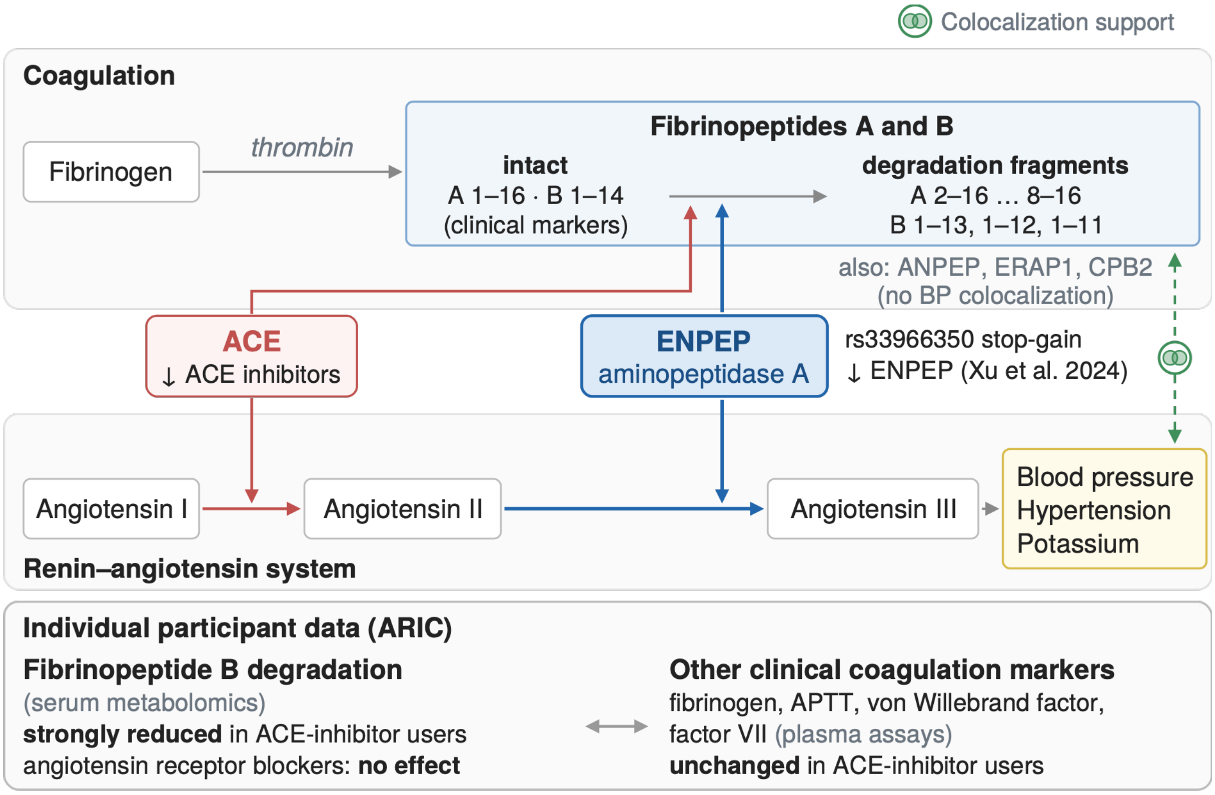

**Extended Data Figure 12: Details on the *SLC16A9* locus.**

**Schematic of known and newly identified relationships at the *SLC16A9* locus.** Gray boxes indicate previously reported associations, and orange boxes and arrows indicate relationships reported in this study. The upward arrow indicates higher metabolite levels with the lead allele. Icons on arrows mark support from colocalization, with the maximum PP.H4 across metabolites, or from MR. Green icons indicate results consistent with prior literature, orange icons indicate results new in this study, and the dashed icon indicates support only with evidence of horizontal pleiotropy.

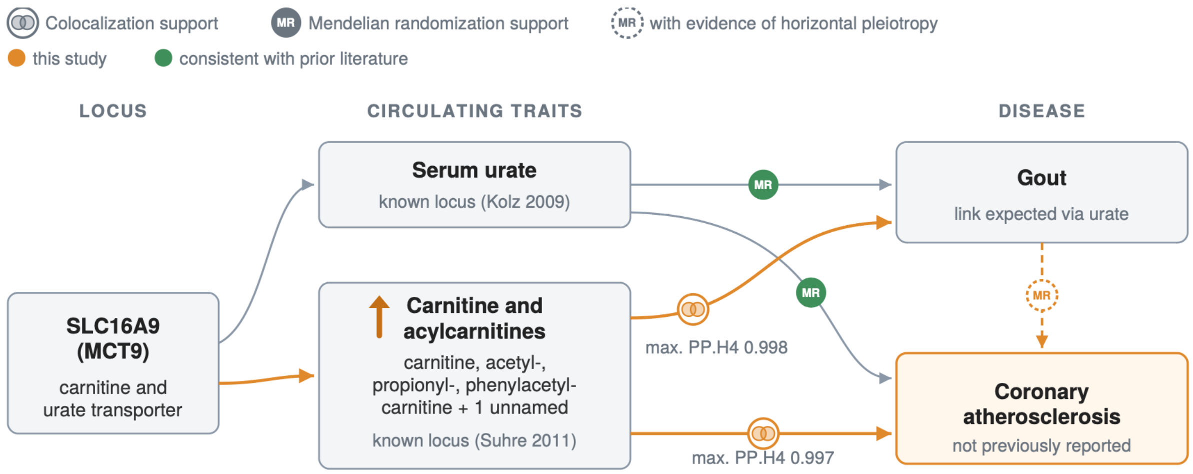

**Supplementary Data 1: Regional association plots for fine-mapped mQTL loci.**

Each panel shows a four-track regional association plot for the fine-mapped region. From top to bottom: **(1)** Posterior Inclusion Probabilities (PIPs) from MeSuSiE multi-ancestry fine-mapping. Variants belonging to credible sets are colored by credible set membership (L1, L2, etc.); remaining variants are shown in gray. The EA index variant is indicated by a purple circle and the AA index variant by an orange triangle. **(2, 3)** Regional association in the European- and African-American-ancestry populations, respectively, showing −log10(*P*) derived from MeSuSiE *Z*-scores. Points are colored by $r^2$ with the corresponding ancestry's index variant (red = high LD, blue = low LD, gray = no LD information). **(4)** Gene models for the locus. The plot title reports information of the metabolite, fine-mapped region and the respective mQTL category (locus-level: EA-specific, AA-specific, or Shared).

**Supplementary Data 2: AlphaGenome Annotation.**

Full list of individual track data from AlphaGenome used to collapse into **Supplementary Tables 22 and 24**.

**Supplementary Note 1: Summary of AlphaGenome predictions for finemapped variants**

Variant effects were summarized using quantile scores, where values exceeding the 99th percentile of the genome-wide score distribution (|quantile| > 0.99) were considered as high-confidence. Of 759 unique finemapped variants across serum and urine, 746 (98.3%) showed at least one high-confidence track across tissues and selected assays. The overall maximum quantile score was most frequently driven by RNA-seq gene expression predictions (658 variants, 88.2%), reflecting the transcriptional impact of the regulatory variants. Among the 363 variants with high-confidence scores in non-expression assays, the strongest predicted effect was most often in histone modifications (ChIP-histone, 39.1%) or transcription factor binding (ChIP-TF, 38.8%), followed by transcription initiation (CAGE, 9.9%) and chromatin accessibility (DNase, 9.4%). At cell-type resolution, the proportion of variants with predicted kidney effects increased further, with 438 variants (57.7%) predicted to regulate kidney cells, including 402 with glomerular and 360 with tubular effects.

**Supplementary Note 2: Medication-based observational analyses.**

Both analyses compared RASi users at Visit 5 with participants taking no antihypertensive medication (Methods). Drug-class-specific comparisons contrasted ACE-inhibitor (ACEi) users and angiotensin-receptor-blocker (ARB) users, respectively, with the same untreated reference group; participants taking other antihypertensive medication without the respective drug class were not included in that comparison. All estimates are from the combined EA+AA sample. Metabolite levels were rank-based inverse-normal transformed, so β is expressed in standard-deviation units; coagulation markers and serum potassium are on their native scales. *P*-values below the double-precision floating-point limit (approximately $10^{-308}$) were computed on the log scale from the regression t-statistic and its residual degrees of freedom (n − 10). For the 13 fibrinopeptide fragments, $P < 0.05/13 = 3.8\times10^{-3}$ (Bonferroni) was considered significant.

Coagulation markers were not measured at Visit 5. The first analysis therefore related medication status at Visit 5 (2011–13) to the seven-marker coagulation panel measured at Visit 1 (1987–89) and to fibrinogen and factor VII measured at Visit 3 (1993–95). Because these measurements precede medication ascertainment by about two decades, this analysis compares the earlier coagulation profile of later RASi users with that of participants who remained untreated, and does not capture a concurrent effect of treatment on these markers. None of the seven coagulation markers measured at Visit 1 was associated with RASi use (lowest *P* = 0.069, protein C), ACEi use (lowest *P* = 0.24) or ARB use (lowest *P* = 0.11). At Visit 3, fibrinogen (β = 9.78, *P* = 0.36) and factor VII (β = −1.60, *P* = 0.68) were likewise not associated with RASi use, although only 306 participants had both measurements and complete covariate data.

*Coagulation markers at Visit 1 by exposure group, each compared with untreated participants (β (SE) in native assay units). n = 4,122–4,209 (RASi), 3,111–3,177 (ACEi) and 2,404–2,455 (ARB) per marker.*

| Outcome | RASi β (SE) | RASi *P* | ACEi β (SE) | ACEi *P* | ARB β (SE) | ARB *P* |
|---|---|---|---|---|---|---|
| **Fibrinogen** | −1.91 (2.03) | 0.35 | −2.12 (2.18) | 0.33 | −0.497 (2.74) | 0.86 |
| **APTT** | −0.00910 (0.108) | 0.93 | −0.0327 (0.121) | 0.79 | 0.0796 (0.137) | 0.56 |
| **von Willebrand factor** | −0.0208 (1.51) | 0.99 | 0.312 (1.64) | 0.85 | −0.537 (1.98) | 0.79 |
| **Factor VII** | 0.749 (0.989) | 0.45 | −0.263 (1.08) | 0.81 | 1.22 (1.28) | 0.34 |

| | | | | | | |
|---|---|---|---|---|---|---|
| **Antithrombin III** | −0.838 (0.770) | 0.28 | −0.992 (0.854) | 0.25 | −0.341 (1.03) | 0.74 |
| **Protein C** | 0.0388 (0.0213) | 0.069 | 0.0278 (0.0237) | 0.24 | 0.0450 (0.0279) | 0.11 |
| **Factor VIII:C** | −0.0692 (1.22) | 0.95 | 0.310 (1.37) | 0.82 | 0.177 (1.59) | 0.91 |

ACEi use was associated with six of the 13 fibrinopeptide fragments at the Bonferroni threshold: higher FPB(1-13) (β = 1.33, $P$ = 2.4×10$^{-296}$), FPA(8-16) (β = 0.52, $P$ = 7.0×10$^{-35}$) and FPA(7-16) (β = 0.45, $P$ = 5.7×10$^{-27}$), and lower FPB(1-12) (β = −1.54, $P$ = 1.2×10$^{-419}$), FPB(1-11) (β = −1.43, $P$ = 1.8×10$^{-374}$) and FPB(1-9) (β = −0.20, $P$ = 1.1×10$^{-6}$). ARB use was not associated with any of these six fragments (all $P$ ≥ 0.097) and with none of the 13 fragments at the Bonferroni threshold; its only nominal association, with FPA(3-16) (β = −0.13, $P$ = 0.0088), involved a fragment not associated with ACEi use ($P$ = 0.69). Estimates for any RASi use lay between those for the two drug classes (e.g. FPB(1-12), β = −1.00, $P$ = 1.1×10$^{-155}$), consistent with dilution of the ACEi effect by ARB users.

Serum potassium was lower with both ACEi use (β = −0.057, $P$ = 1.7×10$^{-5}$) and ARB use (β = −0.096, $P$ = 2.1×10$^{-10}$), so this association is not ACE-specific. Aspartate, glycylvaline and aspartylaspartate, measured in serum and urine at Visit 5, were not associated with RASi, ACEi or ARB use (lowest $P$ = 0.064, glycylvaline in serum, ARB).

*Fibrinopeptide fragments, related metabolites and serum potassium at Visit 5 by exposure group, each compared with untreated participants (β (SE) in standard-deviation units for metabolites, native units for potassium). n = 3,615 (RASi), 2,742 (ACEi) and 2,103 (ARB) for serum metabolites; 1,074, 751 and 548 for urine metabolites; 4,256, 3,207 and 2,480 for potassium. FPA/FPB, fibrinopeptide A/B.*

| **Outcome** | **RASi β (SE)** | **RASi *P*** | **ACEi β (SE)** | **ACEi *P*** | **ARB β (SE)** | **ARB *P*** |
|---|---|---|---|---|---|---|
| **FPB(1-13)** | 0.849 (0.0358) | 7.6×10$^{-116}$ | 1.33 (0.0318) | 2.4×10$^{-296}$ | 0.0136 (0.0406) | 0.74 |
| **FPB(1-12)** | −1.00 (0.0358) | 1.1×10$^{-155}$ | −1.54 (0.0292) | 1.2×10$^{-419}$ | −0.0308 (0.0378) | 0.42 |
| **FPB(1-11)** | −0.933 (0.0351) | 1.3×10$^{-142}$ | −1.43 (0.0294) | 1.8×10$^{-374}$ | −0.0613 (0.0370) | 0.097 |
| **FPB(1-9)** | −0.121 (0.0380) | 0.0015 | −0.205 (0.0419) | 1.1×10$^{-6}$ | 0.0110 (0.0497) | 0.83 |
| **FPA(8-16)** | 0.360 (0.0380) | 5.5×10$^{-21}$ | 0.522 (0.0418) | 7.0×10$^{-35}$ | 0.0588 (0.0472) | 0.21 |
| **FPA(7-16)** | 0.303 (0.0377) | 1.3×10$^{-15}$ | 0.450 (0.0414) | 5.7×10$^{-27}$ | 0.0470 (0.0473) | 0.32 |

| | | | | | | |
|---|---|---|---|---|---|---|
| **FPA(5-16)** | 0.0520 (0.0378) | 0.17 | 0.0881 (0.0413) | 0.033 | −0.0244 (0.0499) | 0.63 |
| **FPA(3-16)** | −0.0325 (0.0378) | 0.39 | 0.0166 (0.0415) | 0.69 | −0.132 (0.0502) | 0.0088 |
| **FPA(2-15)** | −0.0655 (0.0374) | 0.080 | −0.0670 (0.0408) | 0.10 | −0.0485 (0.0494) | 0.33 |
| **FPA(3-15)** | −0.0774 (0.0383) | 0.043 | −0.0848 (0.0420) | 0.043 | −0.0726 (0.0506) | 0.15 |
| **FPA(4-15)** | −0.0273 (0.0380) | 0.47 | −0.0447 (0.0420) | 0.29 | 0.0197 (0.0498) | 0.69 |
| **FPA, des-Ala(1)** | −0.0612 (0.0366) | 0.095 | −0.0512 (0.0399) | 0.20 | −0.0712 (0.0491) | 0.15 |
| **FPA, phospho-Ser(3)** | 0.0429 (0.0365) | 0.24 | 0.0551 (0.0402) | 0.17 | 0.0367 (0.0482) | 0.45 |
| **Aspartate (serum)** | −0.0289 (0.0365) | 0.43 | −0.0580 (0.0400) | 0.15 | 0.0151 (0.0490) | 0.76 |
| **Glycylvaline (serum)** | −0.0101 (0.0347) | 0.77 | −0.0588 (0.0385) | 0.13 | 0.0852 (0.0459) | 0.064 |
| **Aspartate (urine)** | −0.0352 (0.0739) | 0.63 | −0.0498 (0.0792) | 0.53 | −0.0224 (0.0891) | 0.80 |
| **Glycylvaline (urine)** | 0.0271 (0.0805) | 0.74 | −0.0255 (0.0861) | 0.77 | 0.102 (0.103) | 0.32 |
| **Aspartylaspartate (urine)** | −0.0392 (0.0770) | 0.61 | −0.107 (0.0864) | 0.21 | 0.0652 (0.0943) | 0.49 |
| **Potassium (serum)** | −0.0670 (0.0121) | $3.5\times10^{-8}$ | −0.0568 (0.0132) | $1.7\times10^{-5}$ | −0.0956 (0.0150) | $2.1\times10^{-10}$ |

**Supplementary Note 3: Mendelian Randomization regarding *SLC16A9*.**

Of the two exposures tested against gout and CAD, urate was robustly associated with both gout (positive control, β = 1.52, 207 SNPs, $P = 3.4\times10^{-183}$) and CAD (β = 0.103, 199 SNPs, $P = 3.5\times10^{-4}$); for urate→CAD, Cochran's Q indicated heterogeneity ($P = 3\times10^{-16}$, expected for a polygenic instrument) but the MR-Egger intercept showed no directional pleiotropy (*P* = 0.21), supporting a credible causal estimate.

A genome-wide carnitine instrument was not robustly associated with CAD (carnitine, Shin et al. instrument, 126 SNPs, IVW *P* = 0.078, MR-Egger opposite-signed, *P* = 0.71), indicating the carnitine- and urate-transport signals are not statistically separable at this locus.

Each of the five acylcarnitines was tested individually against CAD via both ARIC and an independent external instrument (Shin et al. 2014) where available; none showed a robust association (all IVW $P \geq 0.078$ for the best-powered instrument). One nominal ARIC-only signal (propionylcarnitine, IVW *P* = 0.039) did not replicate in the independent Shin instrument (IVW *P* =

0.335, opposite-signed) and did not survive MR-Egger ($P$ = 0.92) or the weighted-median estimator, so it is not treated as a real signal.

*Genome-wide MR of all five SLC16A9-linked acylcarnitines against CAD, via ARIC and the independent Shin et al. 2014 instrument where available.*

| Metabolite | Source | n SNPs | IVW β | IVW SE | IVW *P* | Egger *P* |
|---|---|---|---|---|---|---|
| **Carnitine** | ARIC | 12 | 0.018 | 0.023 | 0.444 | 0.412 |
| **Carnitine** | Shin2014 | 126 | 0.320 | 0.182 | 0.078 | 0.712 |
| **Acetylcarnitine** | ARIC | 10 | −0.004 | 0.026 | 0.871 | 0.878 |
| **Acetylcarnitine** | Shin2014 | 8 | 0.058 | 0.284 | 0.837 | 0.367 |
| **Propionylcarnitine** | ARIC | 19 | 0.040 | 0.019 | 0.039 | 0.921 |
| **Propionylcarnitine** | Shin2014 | 24 | −0.153 | 0.159 | 0.335 | 0.659 |
| **Phenylacetylcarnitine** | ARIC | 11 | −0.011 | 0.026 | 0.681 | 0.349 |
| **Butyrylcarnitine** | ARIC | 16 | 0.005 | 0.011 | 0.658 | 0.632 |
| **Butyrylcarnitine** | Shin2014 | 83 | −0.019 | 0.038 | 0.608 | 0.395 |

A reverse-direction MR analysis (testing CAD as the exposure) found no evidence that CAD affects urate (84 SNPs, $P$ = 0.095) or any of the five carnitine metabolites (minimum $P$ = 0.081, propionylcarnitine, strict tier; all others higher), arguing against reverse causation. A multivariable MR of carnitine plus triglycerides on CAD was attempted but underpowered (≤ 1 SNP survived three-way alignment across both exposures and the outcome).

A locus-restricted analysis using only SLC16A9 variants as instruments showed the same null pattern against CAD for all five acylcarnitines (single-SNP Wald-ratio $P$ = 0.33–0.37; the Shin2014 three-SNP version $P$ = 0.97), while every one of the five was strongly associated with gout at the same locus ($P = 6\times10^{-15}$ to $10^{-20}$), reinforcing that the locus's CAD-relevant signal runs through urate/gout rather than carnitine transport.

In addition to the continuous-urate instrument above, a genome-wide gout instrument (case-control gout diagnosis as exposure) was associated with CAD (CARDIoGRAMplusC4D: β = 0.042, 83 SNPs, $P = 3.0\times10^{-3}$). However, both Cochran's Q ($P = 2.25\times10^{-4}$) and the MR-Egger intercept ($P$ = $6.7\times10^{-5}$) were significant, indicating genuine directional pleiotropy rather than a clean causal estimate.
